# COVID-19 neutralizing antibodies predict disease severity and survival

**DOI:** 10.1101/2020.10.15.20213512

**Authors:** Wilfredo F. Garcia-Beltran, Evan C. Lam, Michael G. Astudillo, Diane Yang, Tyler E. Miller, Jared Feldman, Blake M. Hauser, Timothy M. Caradonna, Kiera L. Clayton, Adam D. Nitido, Mandakolathur R. Murali, Galit Alter, Richelle C. Charles, Anand Dighe, John A. Branda, Jochen K. Lennerz, Daniel Lingwood, Aaron G. Schmidt, A. John Iafrate, Alejandro B. Balazs

**Author notes:** These authors contributed equally (Wilfredo F. Garcia-Beltran, Evan C. Lam, Michael G. Astudillo, Diane Yang).

## Abstract

COVID-19 exhibits variable symptom severity ranging from asymptomatic to life-threatening, yet the relationship between severity and the humoral immune response is poorly understood. We examined antibody responses in 113 COVID-19 patients and found that severe cases resulting in intubation or death exhibited increased inflammatory markers, lymphopenia, and high anti-RBD antibody levels. While anti-RBD IgG levels generally correlated with neutralization titer, quantitation of neutralization potency revealed that high potency was a predictor of survival. In addition to neutralization of wild-type SARS-CoV-2, patient sera were also able to neutralize the recently emerged SARS-CoV-2 mutant D614G, suggesting protection from reinfection by this strain. However, SARS-CoV-2 sera was unable to cross-neutralize a highly-homologous pre-emergent bat coronavirus, WIV1-CoV, that has not yet crossed the species barrier. These results highlight the importance of neutralizing humoral immunity on disease progression and the need to develop broadly protective interventions to prevent future coronavirus pandemics.

## INTRODUCTION

Coronavirus infectious disease of 2019 (COVID-19), caused by infection with severe acute respiratory syndrome coronavirus 2 (SARS-CoV-2), exhibits significant variability in the severity of presentation. The impact of this variability on the development of protective immune responses and the role of antibodies in disease progression is unclear. There is currently no standard treatment regimen for either mild or severe cases of COVID-19, and there is limited understanding of the impact that current investigational therapies have on immune responses against SARS-CoV-2.

Non-human primates (NHP) that have been exposed to SARS-CoV-2 have been found to develop potent antibody responses and are largely immune to reinfection (Chandrashekar *et al*., 2020; Deng *et al*., 2020). Similarly, animal models testing candidate vaccine approaches have demonstrated that protection against SARS-CoV-2 challenge is positively correlated with the development of high titers of neutralizing antibodies (Mercado *et al*., 2020; Yu *et al*., 2020). Importantly, passive transfer of convalescent sera has been shown to prevent infection in otherwise naive animals, highlighting the crucial role of antibodies in mediating protection against viral infection (Hassan *et al*., 2020; Rogers *et al*., 2020).

In contrast, the role of antibodies on the clearance of established SARS-CoV-2 infection and clinical outcomes is less clear. Ordinarily, infections with viruses require cell-mediated immunity for viral clearance. Antibodies mediate functions such as antibody-dependent cellular cytotoxicity (ADCC) and phagocytosis (ADCP) via innate immune cells such as NK cells and macrophages. Yet, the need for antibodies in the clearance of SARS-CoV-2 infection has been challenged by two recent cases of patients with X-linked agammaglobulinemia who acquired and survived SARS-CoV-2 infection without requiring oxygen or intensive care (Soresina *et al*., 2020). Some studies even propose the possibility of a pathogenic role of antibodies in primary infection via antibody dependent enhancement (ADE) and augmentation of inflammation (Liu *et al*., 2019), although it is believed that this is insufficient to explain the prevalence of severe cases of SARS-CoV-2 infection (Arvin *et al*., 2020). As such, a beneficial, neutral, or harmful role of antibodies in active coronavirus infection remains controversial.

Despite numerous clinical studies presently in progress, no broadly effective standard-of-care treatment has yet emerged for COVID-19. Remdesivir, a nucleotide analog active against SARS-CoV-2, has shown modest benefit in severe COVID-19 cases by improving time to recovery (Beigel *et al*., 2020; Wang *et al*., 2020). Hydroxychloroquine was initially tested in patients based on *in vitro* studies (Wang *et al*., 2020; Z. Chen *et al*., 2020), but subsequent meta-analyses and randomized controlled trials have demonstrated no benefit in preventing or treating COVID-19 (Boulware *et al*., 2020; Tang *et al*., 2020; Ullah *et al*., 2020). Morbidity and mortality due to COVID-19 is largely a consequence of adult respiratory distress syndrome (ARDS) caused by a combination of both hyperinflammatory and hypercoagulable states (Domingo *et al*., 2020). Among experimental treatments currently being evaluated, dexamethasone and other corticosteroids that result in immunosuppression have been shown to reduce disease severity (Siemieniuk *et al*., 2020) and improve survival (Horby *et al*., 2020). Given the involvement of immune dysregulation in the pathology of infection, the consequence of current interventions on the development of humoral immunity is not known.

Recent studies have demonstrated the emergence of SARS-CoV-2 variants containing amino acid substitutions in the viral spike protein targeted by antibodies, raising concerns for potential resistance to neutralization. One mutation, D614G, has rapidly become the predominant transmitted variant by outcompeting wildtype infections (Korber *et al*., 2020). While it has been suggested that this mutant results in a more fit virus, the serological consequences of this change are unclear. Additionally, recent studies in bats have described a novel coronavirus (WIV1-CoV) with high homology to SARS-CoV-2 that uses the same ACE2 receptor for cell entry (Menachery *et al*., 2016). It has been postulated that this virus may present a similar pandemic risk if it were to spread from bats to humans. However, the consequence of prior SARS-CoV-2 seroconversion on neutralization of related coronaviruses like WIV1-CoV has not been described.

In this study, we characterized humoral immune responses and clinical outcomes in 113 SARS-CoV-2-infected patients of varying severity who received a range of treatments, as well as 1,257 pre-pandemic individuals. Our COVID-19 patient cohort contained a wide range of outcomes, including non-hospitalized, hospitalized, intubated, and deceased individuals. We assessed inflammatory markers, IL-6 levels, lymphocyte counts, and demographic variables such as age and sex. A quantitative ELISA that measures IgG, IgM, and IgA antibodies to the receptor binding domain (RBD) of SARS-CoV-2 and a high-throughput neutralization assay using lentiviral vectors pseudotyped with SARS-CoV-2 and WIV1-CoV were developed to assess neutralization potency and cross-neutralizing responses. Remarkably, we find that anti-RBD antibody levels, neutralization titer, and neutralization potency index predicted disease severity and survival, yet lacked cross-neutralizing activity to pre-emergent WIV1-CoV.

Taken together, our results highlight the impact of an effective humoral immune response on COVID-19, as quantified by a neutralization potency index, and describe the influence of current experimental therapies on antibody development. The limited cross-neutralizing potential of antibodies from SARS-CoV-2-infected patients highlights the need to focus future effort on the development of broadly protective interventions to mitigate future coronavirus pandemics.

## RESULTS

### Spectrum of clinical severity of SARS-CoV-2 infection

A cross-sectional cohort of 113 COVID-19 cases confirmed by SARS-CoV-2 nasopharyngeal PCR was studied and followed for at least 3 months. The cohort was divided into the following five groups based on disease severity, outcomes, and pre-existing health status: (*i*) non-hospitalized, which were never admitted to the hospital due to COVID-19; (*ii*) hospitalized, which were admitted for at least one day but were never intubated and were eventually discharged, (*iii*) intubated, which were intubated for at least one day but were subsequently extubated and discharged; (*iv*) deceased, which passed away due to COVID-19; and (*v*) immunosuppressed, which were includes some non-hospitalized, hospitalized, and intubated patients, but none deceased) (**Supplementary Table 1**). When compared to non-hospitalized individuals, all cases of COVID-19 resulting in hospital admission were significantly older in age (median age 63 versus 28, *p* < 0.0001) and there was a significant enrichment for males in severe cases resulting in intubation and/or death (74% versus 51% males, *p* = 0.02) (**Figure 1A**), consistent with prior studies (Meng *et al*., 2020; N. Chen *et al*., 2020). Laboratory data showed that clinical severity correlated with markers of inflammation, namely, peak serum levels of C-reactive protein (**Figure 1B**), ferritin (**Figure S1A**), D-dimer (**Figure S1B**), lactate dehydrogenase (**Figure S1C**), and IL-6 (**Figure 1C**), as well as lymphopenia (**Figure 1D**), as has been previously shown (Wang, 2020; Wynants *et al*., 2020; X. Chen *et al*., 2020; Zhou *et al*., 2020). Interestingly, COVID-19 severity was also associated with peak serum levels of troponin-T (**Figure S1D**), a marker of myocardial damage and/or ischemia that may reflect cardiac injury, as has been previously described (Tersalvi *et al*., 2020). Altogether, our cohort contained a wide range of clinical presentations of SARS-CoV-2 infection with our analyses confirming previously described associations.

**Figure 1:**
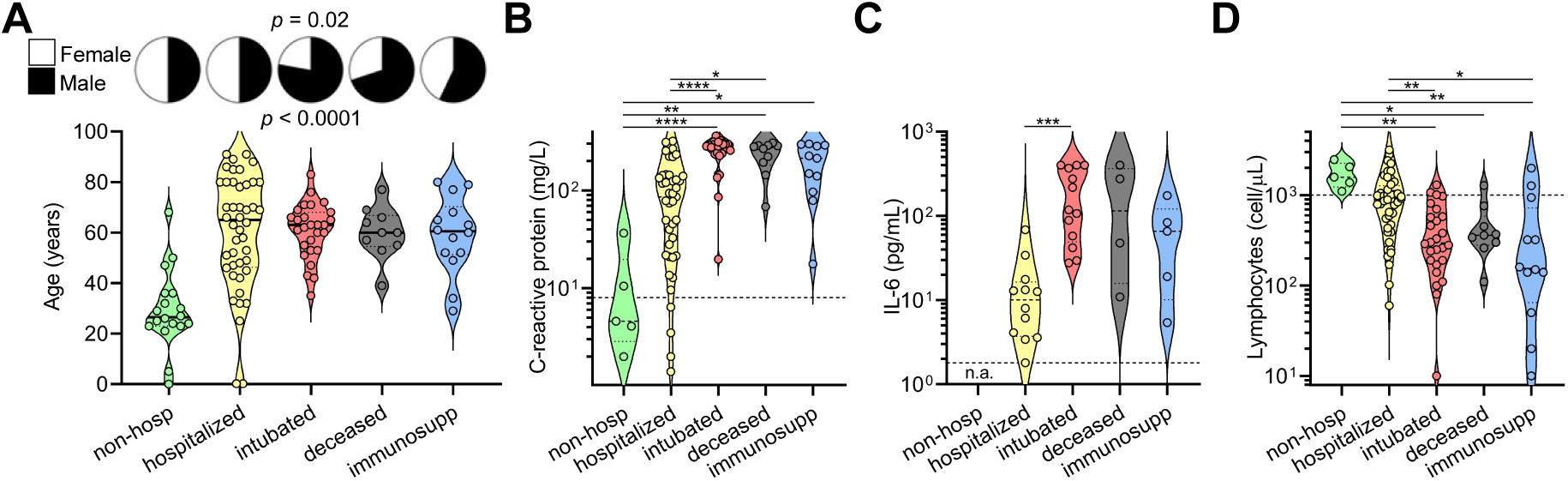
Clinical severity of SARS-CoV-2 infection is influenced by patient characteristics and coupled to clinical laboratory data. (**A**) A cross-sectional cohort of COVID-19 patients (*n* = 113) was divided into groups of varying clinical severity, i.e., non-hospitalized (*n* = 18), hospitalized (*n* = 45), intubated (*n* = 27), deceased (*n* = 10), and immunosuppressed (*n* = 13) and analyzed for their age and sex. Median age was 28 years in patients who were never hospitalized (*n* = 20; includes 2 from immunosuppressed group) and 63 years in all patients who were admitted to the hospital (*n* = 93), with statistical significance of *p* < 0.0001 with *t* test. Fisher’s exact test on the number of males who were intubated or deceased (*n* = 31 males out of 42 total; includes 5 from immunosuppressed group who were intubated) versus not (*n* = 36 males out of 71 total) demonstrated a significant enrichment with *p* = 0.02. (**B-D**) Peak levels of C-reactive protein and IL-6 as well as lymphocyte count nadir are presented in violin plots when data was available. In **C**, none of the non-hospitalized patients had serum IL-6 levels measured (n.a., not assessed). For **B** and **C**, clinical laboratory-defined cut-offs of the upper limit of normal are indicated with a dotted line; for **D**, the dotted line represents the lower limit of normal. For each parameter, a non-parametric ANOVA was performed; statistical significance is indicated with the following notations: **** *p* < 0.0001, *** *p* < 0.001, ** *p* < 0.01, and * *p* < 0.05.

### Quantitative SARS-CoV-2 receptor binding domain IgG, IgM, and IgA ELISA

An ELISA that quantitatively measures IgG, IgM, and IgA antibodies that target the receptor binding domain (RBD) of SARS-CoV-2 spike protein was developed to characterize humoral immune responses (**Figure S2A**), similar to what we have previously described (Iyer *et al*., 2020; Roy *et al*., 2020). Quantitation was achieved by using a standard curve consisting of purified IgG, IgM, and IgA isotype of a monoclonal antibody, CR3022 (**Figure 2A**), that cross-reacts to bind both SARS-CoV and SARS-CoV-2 RBD (**Figure S2B**) (ter Meulen *et al*., 2006; Tian *et al*., 2020). RBD was chosen over full-length spike as the target protein because of its specificity to SARS-CoV-2 as well as its ease of production and stability (Stadlbauer *et al*., 2020). Full-length spike has more regions of homology to other coronaviruses that may cause greater false positivity, as has been shown between SARS-CoV, SARS-CoV-2, MERS-CoV, and common cold CoVs (Chan *et al*., 2005; Ju *et al*., 2020). In addition, studies have shown that RBD is the main target of coronavirus neutralizing antibodies (He *et al*., 2005).

**Figure 2:**
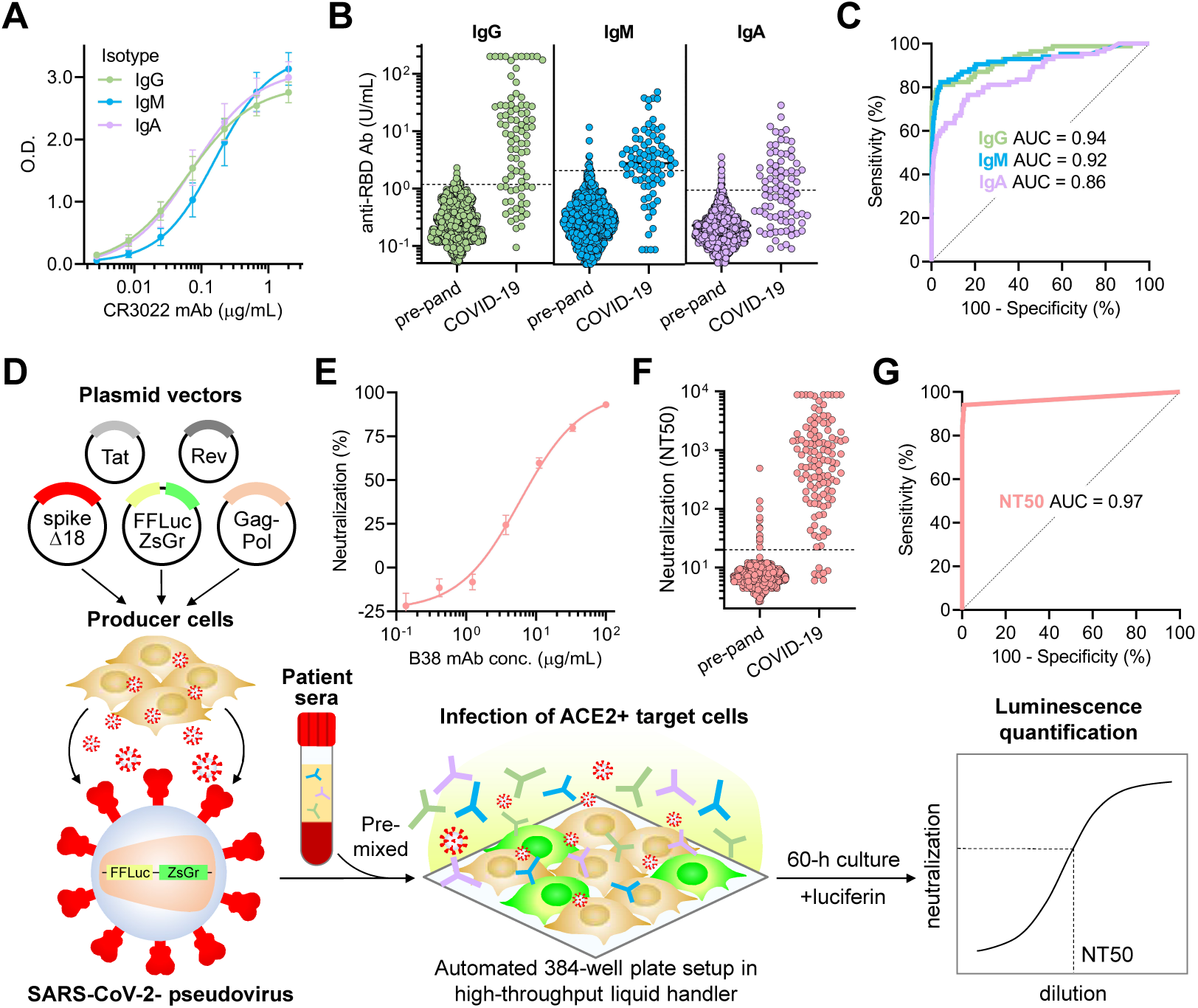
Quantitative SARS-CoV-2 receptor binding domain ELISA and high-throughput SARS-CoV-2 pseudovirus neutralization assay reveal highly variable IgG, IgM, and IgA responses and neutralization potency after SARS-CoV-2 infection. **(A)** For quantitation of anti-RBD IgG, IgM, and IgA antibodies, a 7-point standard curve consisting of a SARS-CoV and -CoV-2 RBD-binding monoclonal antibody, CR3022, in all three isotypes (CR3022-IgG, CR3022-IgM, and CR3022-IgA) was used as a reference to interpolate O.D. values from samples and calculate units/mL (U/mL), with 1 U/mL defined as the equivalent reactivity caused by 1 μg/mL of the corresponding CR3022 monoclonal antibody **(B)** Anti-RBD IgG, IgM, and IgA antibodies were measured in both pre-pandemic samples (*n* = 1,257) and COVID-19 patient samples (*n* = 85). Dotted lines indicate the threshold of seropositivity that achieves >99.0% specificity on ROC analyses. **(C)** ROC analyses for each assay were done to assess how seropositivity predicted COVID-19 status. Area under the curve (AUC) was 0.94 for IgG, 0.92 for IgM, and 0.86 for IgA. Cut-offs of 1.18 U/mL for IgG achieved a sensitivity of 73%, 2.14 U/mL for IgM achieved 66%, and 0.95 U/mL for IgA achieved 48%, with >99.0% specificity for all three. **(D)** A schematic of the high-throughput SARS-CoV-2 pseudovirus neutralization assay is shown. **(E)** Validation of the neutralization assay using a recently discovered neutralizing monoclonal antibody, B38, was performed and showed an IC50 of 6 μg/mL. **(F)** Neutralization titers that achieved 50% neutralization (NT50) were calculated for pre-pandemic samples (*n* = 1,220, individuals on antiretroviral therapy excluded) and COVID-19 patient samples (*n* = 118). **(G)** An ROC analysis demonstrated an AUC of 0.97, with an NT50 cut-off of 20 achieving sensitivity of 94% and specificity of >99.0%.

We determined the sensitivity and specificity of this assay by assessing anti-RBD antibody levels in a cohort of SARS-CoV-2-infected patient serum samples collected between 14 to 42 days after symptom onset (*n* = 85) in order to maximize seropositivity for IgG, IgM, and IgA. We also assessed 1,257 pre-pandemic serum samples composed of a large unbiased cohort (*n* = 1,124) and selected cohorts of individuals (*n* = 133) with positive serology results for cytomegalovirus, varicella-zoster virus, hepatitis B virus, hepatitis C virus, HIV, syphilis, Toxoplasma, and/or rheumatoid factor (**Figure 2B**). Anti-RBD IgG, IgM, and IgA levels were measured for each sample by interpolation on to the standard curve and a receiver operating curve (ROC) analysis was used to determined optimal cut-offs that distinguished SARS-CoV-2-infected patients from pre-pandemic controls (**Figure 2C**). Cut-offs of 1.18 U/mL for anti-RBD IgG achieved 73% sensitivity, 2.14 U/mL for anti-RBD IgM achieved 66% sensitivity, and 0.95 U/mL for anti-RBD IgA achieved 48% sensitivity, with >99.0% specificity for all three.

To assess the cross-reactivity of anti-RBD IgG in sera of SARS-CoV-2 seropositive individuals, we modified our ELISA to detect IgG antibodies against the RBD of SARS-CoV and MERS-CoV. Interestingly, no cross-reactivity was seen to SARS-CoV RBD despite 73% homology, nor to MERS-CoV, which has only 17% homology (**Figure S2C** and **S2D**). Additional experiments measuring IgG antibodies against the RBD of two common cold coronaviruses—NL63, which has 20% homology to SARS-CoV-2 RBD, and HKU1, which has 1.9% homology (**Figure S2C**)—showed a seroprevalence of >95% (**Figure S2E**), as has been shown in previously published studies (Gorse *et al*., 2010), with no correlation between the IgG antibody levels of NL3 or HKU1 with SARS-CoV-2 (**Figure S2E**). These data show that anti-RBD IgG antibodies induced during SARS-CoV-2 infection do not cross-react to recognize the RBD of other pandemic coronaviruses. In addition, anti-RBD IgG antibodies to common cold coronaviruses appear to not provide detectable pre-existing reactivity to SARS-CoV-2 RBD nor do they correlate with anti-RBD IgG levels in COVID-19 patients. Overall, these data suggest that natural infection with coronavirus results in anti-RBD antibodies with limited cross-reactivity.

### High-throughput SARS-CoV-2 pseudovirus neutralization assay

Previous studies have demonstrated the potential to pseudotype retroviral vectors with SARS-CoV spike proteins (Moore *et al*., 2004). However, pseudoviruses bearing SARS-CoV-2 spike produced by these methods yield low titers (Nie *et al*., 2020), hampering large-scale testing of neutralization. Recently, a forward genetics approach identified an efficiently replicating vesicular stomatitis virus (VSV) variant encoding SARS-CoV-2 spike containing a truncated form lacking the C-terminal 21 amino acids (Case *et al*., 2020). Interestingly, previous studies also showed a role of the cytoplasmic tail of SARS-CoV in altering surface expression and fusogenic potential (Corver *et al*., 2009). To determine whether analogous truncations might improve SARS-CoV-2 pseudovirus production, we examined the cell-surface expression of truncated forms of SARS-CoV-2 spike and found that removal of 18 amino acids from the C-terminus (Δ18) resulted in significantly greater cell-surface expression and higher titers of pseudovirus (**Figure S2F-H**). This truncation removed a putative ER-retention signal (Lontok, Corse and Machamer, 2004; McBride, Li and Machamer, 2007; Ujike *et al*., 2016) while retaining cysteine-rich domains that are highly conserved among coronaviruses. Using these spike modifications, we developed a CoV pseudovirus neutralization assay compatible with high-throughput liquid handling instrumentation in 384-well plate format using our previously published lentiviral vector system expressing both luminescent and fluorescent marker transgenes (**Figure 2D**) (Crawford *et al*., 2020).

To validate our assay, the potency of a neutralizing monoclonal antibody, B38, and a non-neutralizing monoclonal antibody, CR3022, both of which target SARS-CoV-2 RBD with known IC50 values, was determined. This yielded IC50 values of ∼6 μg/mL for B38 and undetectable (>100 μg/mL) for CR3022, which were in agreement with previous reports (Tian *et al*., 2020; Wu *et al*., 2020) (**Figure 2E** and **S2I**). In addition, we found that luciferase activity was directly proportional to the number of infected (i.e. ZsGreen+) cells, providing flexibility in assay readout (**Figure S2J**). To determine the performance of our assay on human sera, we measured the neutralization potency of human sera from 1,220 pre-pandemic individuals and 118 COVID-19 patient samples >15 days after symptoms onset, with a dilution range of 1:12 to 1:8,748. The dilution titer that achieved 50% neutralization (NT50) was calculated for each specimen and ROC analysis was performed, revealing that an NT50 threshold of 1:20 achieves a sensitivity of 94% and specificity of >99.0% in identifying COVID-19 patients (**Figure 2G-H**). Overall, we found median titers of 1:664 in COVID-19 patients, with potency ranging from <1:12 to >1:8,748. Comparatively, titers of 1:5 for yellow fever vaccination, 1:8 for rubella vaccination, and 1:40 for influenza vaccination are considered to indicate protective immunity (Hannoun, Megas and Piercy, 2004; Plotkin, 2010). Altogether, we established a highly accurate high-throughput SARS-CoV-2 pseudovirus neutralization assay that can accurately quantify the neutralization potency of humoral immune responses directed to SARS-CoV-2 spike protein.

As a separate note for investigators using pseudovirus neutralization assays, we excluded pre-pandemic individuals taking antiretroviral therapy for human immunodeficiency virus infection or pre-exposure prophylaxis (*n* = 37 in the original cohort of 1,257) after finding that potent inhibition of pseudovirus infection occurred in a majority of these individuals (**Figure S2K**). We believe this was due to antiretroviral compounds in the patients’ sera inhibiting transduction with our lentivirus-based vector system, thus generating an artifact. Of note, non-documented antiretroviral use may explain a proportion of the false positives observed in the remaining specimens (*n* = 12 out of 1,220).

### Relationship between neutralizing humoral immunity in SARS-CoV-2 infection and clinical severity

We proceeded to analyze antibody responses in our cohort of 113 COVID-19 patients as well as a negative control cohort of 37 healthy blood donors, and found that in contrast to the typical kinetics of antibody responses in viral infections (i.e. IgM before class-switched IgG and IgA), anti-RBD IgG antibodies appear almost simultaneously with anti-RBD IgM antibodies after symptom onset, and only a subset of individuals generate anti-IgA antibodies concomitantly (**Figure 3A-C**). Interestingly, the development and quantity of anti-RBD IgG antibodies appeared to be increased and sustained in the time frame analyzed (up to 72 days), while anti-RBD IgM and IgA antibodies waned after ∼42 days. Neutralization titers closely resembled anti-RBD IgG levels and were similarly sustained over time (**Figure 3D**). Overall, seropositivity at >14 days after symptom onset was 74% for anti-RBD IgG, 61% for anti-RBD IgM, 41% for anti-RBD IgA, and 89% overall for any antibody. Neutralization was detected in 95% of samples >14 days after symptom onset.

**Figure 3:**
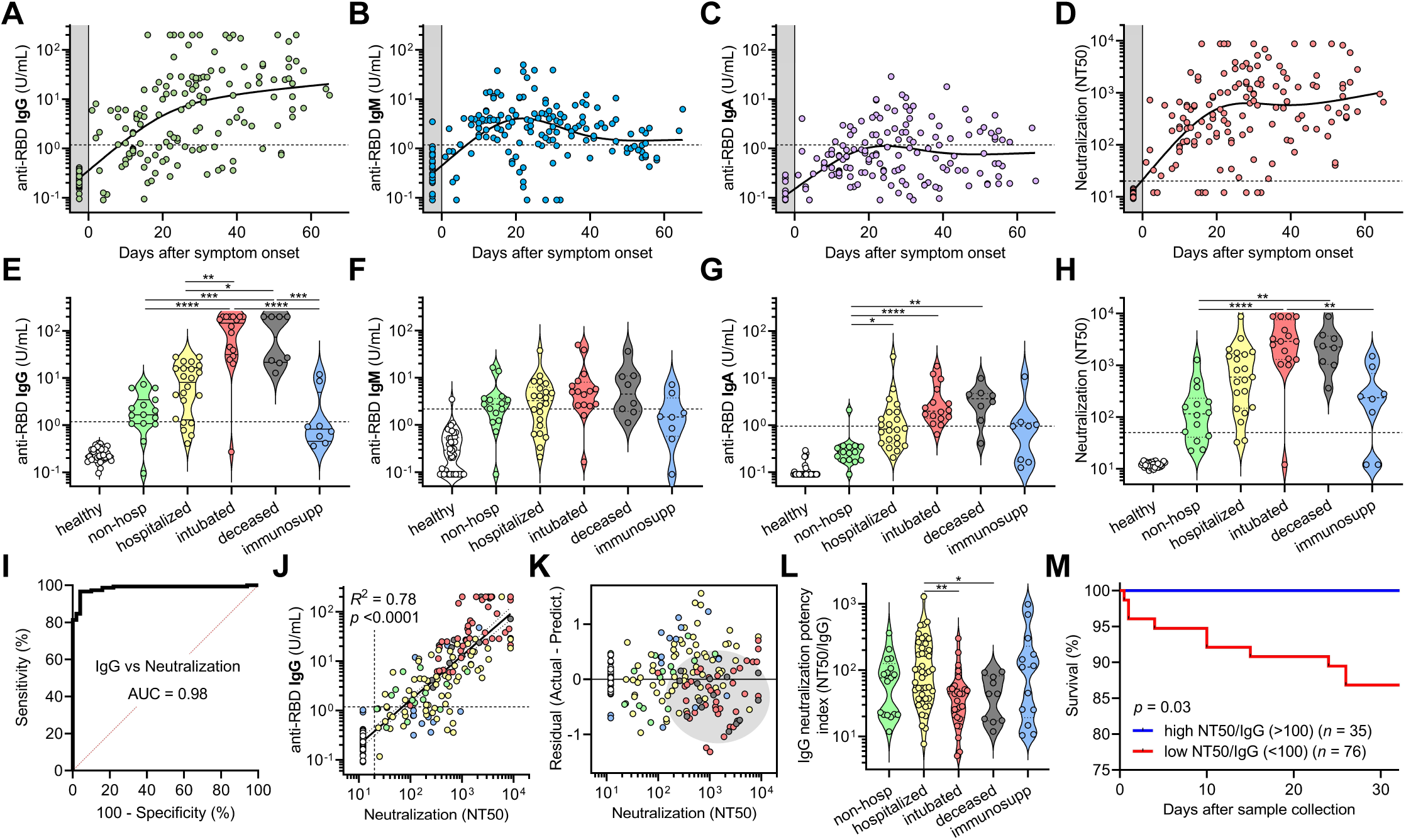
SARS-CoV-2 antibody levels and neutralization potency predict clinical severity and survival. (**A-C**) Anti-RBD IgG, IgM, and IgA levels were plotted over days after symptom onset for confirmed COVID-19 cases for which data of symptom onset was known (*n* = 98 patients, *n* = 147 samples total). Healthy blood donors (*n* = 37) are included as a negative control within the gray region. The dotted lines indicate the cut-offs for anti-RBD IgG, IgM, and IgA seropositivity. (**D**) Titers that achieve 50% neutralization (NT50) were plotted over days after symptom onset for each patient sample. (**E-H**) Patient samples were selected for collection between 14 and 42 days after symptom onset (earliest time point for each patient), and for each cohort of healthy blood donors, non-hospitalized, hospitalized, intubated, deceased, and immunosuppressed patients, anti-RBD IgG, IgM, IgA, and neutralization (NT50) was plotted. Non-parametric multivariate ANOVA was performed for each (excluding healthy blood donors); statistical significance is indicated as follows: **** *p* < 0.0001, *** *p* < 0.001, ** *p* < 0.01, and * *p* < 0.05. (**I-J**) An ROC and log-log regression analyses were performed on IgG versus neutralization. For **J**, the severity cohort is indicated as follows: healthy (white), non-hospitalized (green), hospitalized (yellow), intubated (red), deceased (gray), and immunosuppressed (blue). For **J**, Pearson correlations were performed and *R*^2^ and *p* values are indicated. **(K)** A residual plot for neutralization titer was generated from the log-log correlation. The gray ellipse indicates a cluster of samples from intubated (red) and deceased (gray) patients. **(L)** Neutralization potency index (NT50/IgG) was calculated for all 113 patients (at earliest time point) and plotted by cohort. A non-parametric multivariate ANOVA was performed without correction for multiple comparisons; unadjusted *p* values are indicated as follows: ** *p* < 0.01, * *p* < 0.05. **(M)** Survival analysis of COVID-19 patients classified as having a high (≥100) (*n* = 30) or low (<100) (*n* = 68) neutralization potency index (NT50/IgG) was performed using Kaplan-Meier method and revealed significantly decreased risk of death in low neutralization potency individuals (*p* = 0.03).

To assess the humoral immune response among the pre-defined cohorts of varying disease severity, we focused on patients for which samples were collected between 14 and 42 days after symptom onset (*n* = 85). This time frame was chosen to prevent biases resulting from time of sampling post-infection (**Figure S3A**), which is known to have a significant impact on the magnitude of antibody responses. We found that severely ill patients that were intubated or passed away due to COVID-19 had the highest anti-RBD IgG and IgA levels, but no significant differences were seen for IgM (**Figure 3E-G**). These individuals also had the highest neutralization titers (**Figure 3H**). In contrast, individuals that were not hospitalized had the lowest anti-RBD IgG and IgA levels and neutralization titers. Unsurprisingly, immunosuppressed individuals—none of whom passed away—had significantly blunted IgG, IgA, and neutralizing responses. Upon analyzing anti-RBD antibody seropositivity and neutralization titer, we found IgG seropositivity was an excellent predictor of neutralization with a sensitivity of 78% and specificity of 100% (**Figure 3I**). When seropositivity for any anti-RBD antibody was present, neutralization could be predicted with a sensitivity of 91% and specificity of 94%. Anti-RBD IgG levels correlated the most with neutralization (*R*^2^ = 0.76) (**Figure 3J**), while anti-RBD IgM and IgA exhibited weaker correlations with neutralization titer (*R*^2^ = 0.49 for IgM and *R*^2^ = 0.64 for IgA) (**Figure S3B-C**). Triple positivity for anti-RBD IgG, IgM, and IgA was enriched in severely ill patients and was associated with the highest neutralization titers (**Figure S3D-E**). However, anti-RBD IgM and IgA alone were capable of neutralization in serum samples that were negative for anti-RBD IgG (**Figure S3D**). Altogether, these results highlight anti-RBD IgG as the main contributor to neutralization from patient sera, with a contributory role from anti-RBD IgM and IgA.

Although anti-RBD IgG levels correlated with neutralization by regression analysis, there was variability that appeared to segregate by our pre-defined severity cohorts (**Figure 3J**). To better visualize this, we plotted residuals of each neutralization titer subtracted from its predicted titer based on the regression (**Figure 3K**). This revealed that samples from severely ill patients were biased towards lower-than-predicted neutralization titers, suggesting that they harbored higher levels of non-neutralizing anti-RBD IgG antibodies that did not contribute to neutralization. Consequently, we calculated a neutralization potency index (NT50/IgG) for each patient, and found that intubated or subsequently deceased patients had a significantly lower index (**Figure 3L**), with all deceased patients having an index <100. Accordingly, when patients were classified as having neutralization potency indices that were ‘high’ (≥100) or ‘low’ (<100), there was a significant risk of death in the days following sample collection in the ‘low’ index group (87% 30-day survival, *n* = 68) and there were no deaths in the ‘high’ index group (100% 30-day survival, *n* = 30) (p = 0.03; **Figure 3M**). Of note, this finding was true across our entire cohort of 111 COVID-19 patients (including non-hospitalized and immunosuppressed individuals) for which both anti-RBD IgG. In addition, neutralization potency index did not correlate with days after symptom onset and remained predictive of survival when using a Cox proportional hazards model that accounted for age, sex, hospitalization status, intubation status, and days between symptom onset and sample collection (*p* = 0.03). These results suggest that neutralization potency index may help risk stratify patients irrespective of where they are in their disease course. Altogether, severity of SARS-CoV-2 infection significantly correlates with higher anti-RBD antibody levels but sub-optimal neutralization potency is a significant predictor of mortality.

### The influence of pre-existing medical conditions and COVID-19 therapies on humoral immune responses to SARS-CoV-2

To explore the influence of pre-existing medical conditions and COVID-19 therapies on humoral immune responses to SARS-CoV-2 we performed multivariate analysis of all available demographic, clinical, laboratory, and experimental data (**Figure S4**). With the exception of immunosuppressed individuals, which had significantly decreased antibody and neutralizing responses, our cohort was not large enough to conclusively detect the effects of particular pre-existing medical conditions on the overall humoral immune response. However, a principle components analysis (PCA) that included demographic data, pre-existing medical conditions, laboratory data, treatments received, anti-RBD antibody levels and neutralization titers but not clinical outcomes demonstrated clustering of patients by the severity cohorts (**Figure 4A**). Principal components were mainly influenced by inflammatory markers, anti-RBD antibody levels, and neutralization titers, but a contribution from pre-existing medical conditions such as hypertension and diabetes was observed (**Figure 4B**).

**Figure 4:**
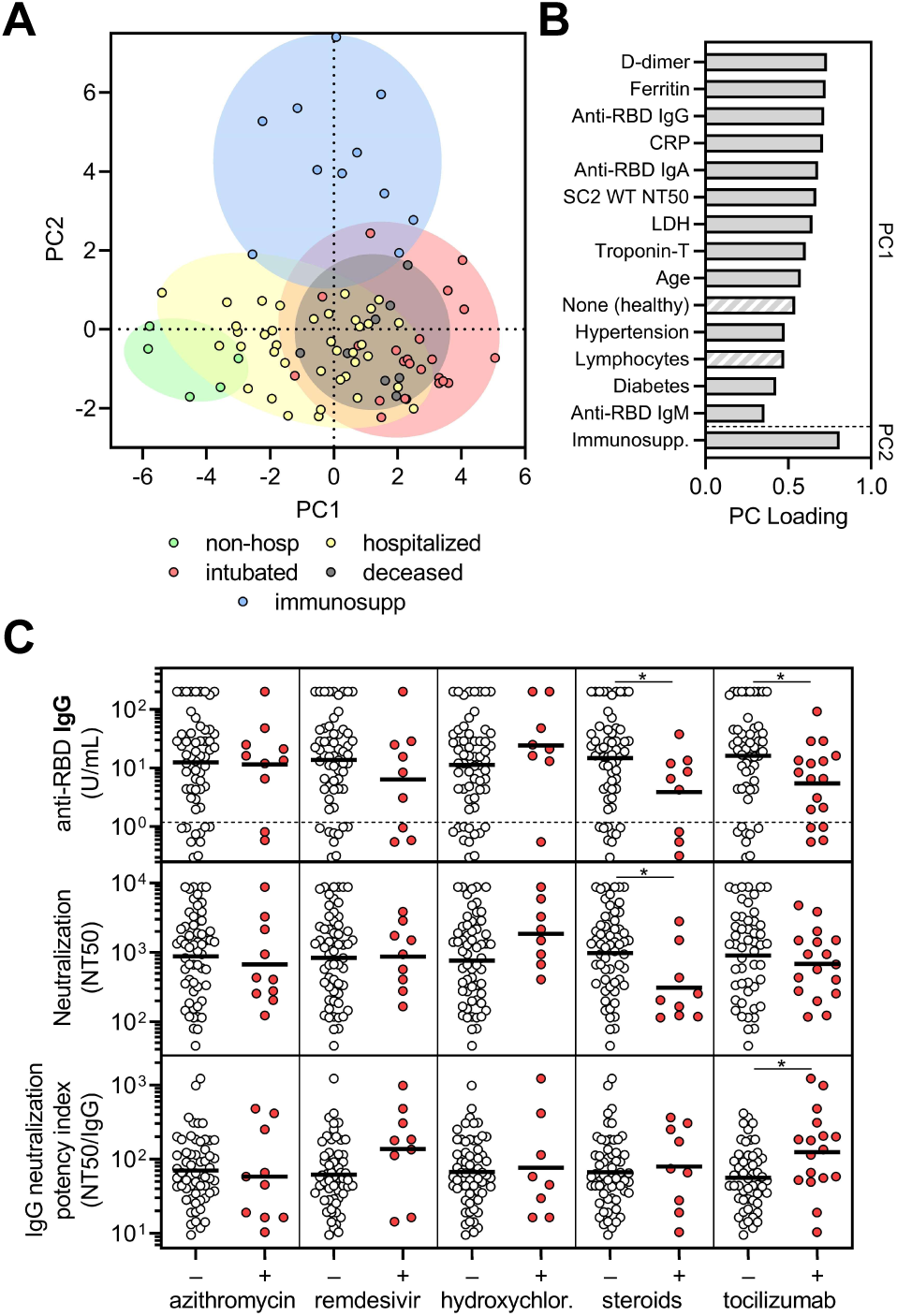
Corticosteroid and tocilizumab therapy decrease humoral immune responses to SARS-CoV-2. **(A)** Principle components analysis was performed using the following variables: age, sex language, pre-existing medical conditions, treatments received, clinical laboratory data (ferritin, CRP, D-dimer, LDH, troponin-T, and lymphocyte nadir), anti-RBD antibody levels, and neutralization titers. The severity cohort of each patient is indicated by color. Patients with missing data were excluded. **(B)** Loading of principle components (PC) is shown. Hatched bars indicate negative loading. **(C)** Sub-analyses on COVID-19 patients that were in the hospital for at least 3 days to (*n* = 69) were performed on the last collected specimen to show the effect of azithromycin (*n* = 10 treated), remdesivir (*n* = 9 treated), hydroxychloroquine (*n* = 8 treated), corticosteroids (*n* = 9 treated), and tocilizumab (*n* = treated as part of a trial with 2:1 randomization to placebo) on anti-RBD IgG levels (upper panel), neutralization titer (middle panel), and neutralization potency index (NT50/IgG) (lower panel). A *t* test was performed for each comparison; * indicates unadjusted *p* < 0.05.

To assess the effect of different treatments on the humoral immune response, we performed an analysis limited to samples collected from patients that had initiated treatment and were in the hospital for at least 3 days (*n* = 69). COVID-19-directed treatment regimens included azithromycin (an antibiotic with anti-inflammatory properties), remdesivir, hydroxychloroquine, corticosteroids, and tocilizumab (an anti-IL-6 receptor antibody). Of note, individuals in the tocilizumab-treated cohort included 3 individuals known to receive tocilizumab for compassionate use and 16 patients enrolled in a blinded clinical trial with 2:1 tocilizumab-to-placebo randomization (i.e. some patients might have received placebo). Azithromycin, remdesivir, and hydroxychloroquine—for which there was concern of attenuating antibody responses (de Miranda Santos and Costa, 2020)—did not significantly affect anti-RBD antibody levels or neutralization titers in our cohort (**Figure 4C**). However, we found that use of corticosteroids and tocilizumab significantly decreased anti-RBD IgG concentration, and in the case of corticosteroids, neutralization titer (**Figure 4C**). Corticosteroids are a general immunosuppressant known to decrease antibody production, whereas IL-6 signaling is important in several aspects of antibody responses (Kopf *et al*., 1998). Interestingly, tocilizumab-treated patients had a significant increase in the neutralization potency index stemming from the larger effect on anti-RBD IgG as compared to neutralization (**Figure 4C**). This result raises new questions regarding the role of IL-6 signaling in the production of non-neutralizing versus neutralizing antibodies and how these might become de-coupled, although a selection bias cannot be excluded. Altogether, immunomodulatory therapies, some of which have shown clinical efficacy or are actively being studied, influence humoral immune responses in SARS-CoV-2-infected patients.

### Cross-neutralization of SARS-CoV-2-infected patients to emerging coronaviruses

Given the importance of humoral immunity in preventing most viral infections, the recent emergence of a mutation in the SARS-CoV-2 spike protein (D614G) has raised concerns for the potential for convalescent patients to become re-infected. Studies have demonstrated that this variant may possess greater replicative fitness and an altered conformation of the spike protein that may render pre-existing immunity less effective (Korber *et al*., 2020). To determine the impact of this variant on the neutralization potency of sera from patients previously infected with SARS-CoV-2, we introduced the D614G mutation into the SARS-CoV-2 Δ18 spike (**Figure 5A**). When characterizing this new construct, we found that both surface expression and infectivity were further increased relative to that of the D614 SARS-CoV-2 Δ18 spike (**Figure 5B** and **S5A**,**C**,**D**,**F**), in line with previous studies (Korber *et al*., 2020). We tested this new pseudovirus, normalizing for infectious units per well, against the same panel of patient samples and found an increase in neutralizing titers that was very small but statistically significant (**Figure 5C-D**), an effect that was seen in a prior study (Korber *et al*., 2020). This indicates that individuals that have been infected with either D614 wild-type or G614 mutant SARS-CoV-2 will have cross-neutralization to the opposite strain, both of which are circulating in Boston, Massachusetts (Lemieux *et al*., 2020) and were likely represented in our study cohort.

**Figure 5:**
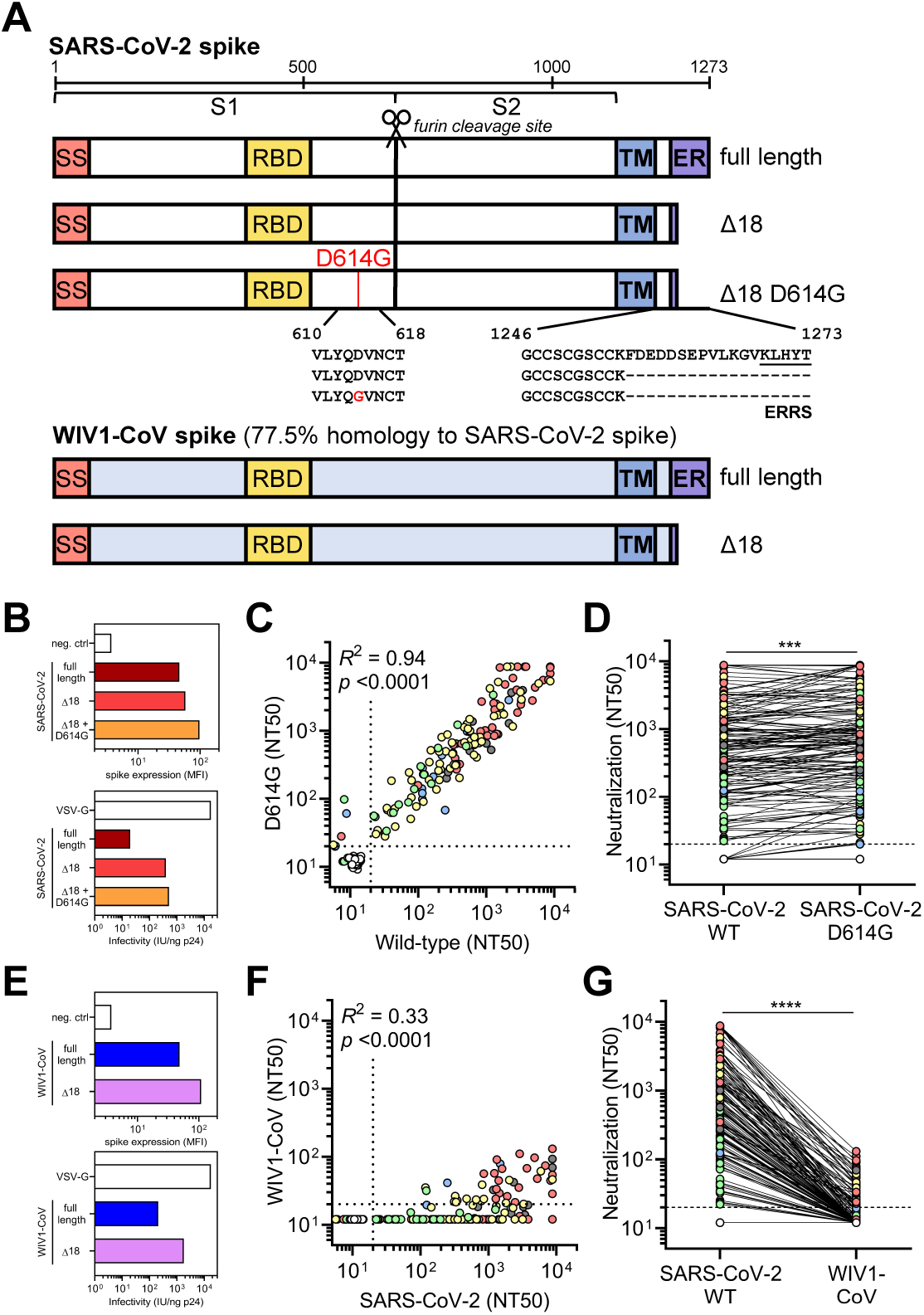
SARS-CoV-2-infected patient sera cross-neutralizes both wild-type and D614G mutant SARS-CoV-2 spike but not the highly homologous pre-emergent bat coronavirus WIV1-CoV. **(A)** A schematic of the SARS-CoV-2 and WIV1-CoV spike proteins, including full-length, truncated (Δ18), and mutant (D614G) forms is shown. Full-length WIV1-CoV spike has 77.5% sequence homology to SARS-CoV-2 spike and has the same putative ER retention signal (ERRS) as SARS-CoV-2. **(B)** Expression of full-length, Δ18, and Δ18 D614G SARS-CoV-2 spike constructs in 293T cells in comparison to empty vector (neg. ctrl) was measured by flow cytometry (upper panel). Infectivity of lentivirus, which was defined as the infectious units divided by the quantity of p24 in lentiviral supernatant, was also measured and compared to VSV-G-pseudotyped lentivirus (lower panel). (**C-D**) Cross-neutralization of serum samples from COVID-19 patients that were non-hospitalized (green, *n* = 16), hospitalized (yellow, *n* = 67), intubated (red, *n* = 43), deceased (gray, *n* = 15), or immunosuppressed (blue, *n* = 21) and healthy blood donors (*n* = 35) was measured for wild-type versus D614G mutant SARS-CoV-2 Δ18 spike pseudovirus. For **C**, Pearson correlations were performed and *R*^2^ and *p* values are indicated; for **D**, paired, non-parametric *t* test was performed; *** indicates *p* < 0.001. (**E**) Similar to **B**, expression and infectivity of full length and Δ18 WIV1-CoV spike was measured. (**F-G**) Similar to **C-D**, cross-neutralization of serum samples from COVID-19 patients was measured for wild-type SARS-CoV-2 versus WIV1-CoV pseudovirus. For **F**, Pearson correlations were performed and *R*^2^ and *p* values are indicated; for **G**, paired, non-parametric *t* test was performed; **** indicates *p* < 0.0001.

The emergence of SARS-CoV, MERS-CoV, and now SARS-CoV-2 within the last two decades has demonstrated the ability of zoonotic coronaviruses to cross the species barrier and pose pandemic threats. This has prompted microbiologists and epidemiologists to seek out and characterize zoonotic coronaviruses that have the potential to cross into humans. Recent studies in bats have identified a novel coronavirus, Wuhan Institute of Virology 1 coronavirus (WIV1-CoV), which, like SARS-CoV-2 and SARS-CoV, has a spike that uses ACE2 receptor for cell entry and bears high sequence homology to both SARS-CoV (92%) and SARS-CoV-2 (77%). We generated WIV1-CoV pseudovirus using an analogous spike truncation (Δ18) (**Figure 5A**), which resulted in high expression of WIV1-CoV spike on producer cells as well as infectivity and titer (**Figure 5E** and **S5B**,**C**,**E**,**F**). These results suggest that this C-terminal truncation can serve as a general approach for modifying coronavirus spike proteins for efficient pseudovirus production. Interestingly, WIV1-CoV spike could be detected at the cell surface by the SARS-CoV and -CoV-2-specific monoclonal antibody CR3022 (**Figure S5B**), a finding that, to the best of our knowledge, has not been previously described. Using WIV1-CoV pseudovirus, we found that sera from SARS-CoV-2-infected individuals showed a lack of cross-neutralization except for relatively low-level neutralization in a few individuals with very high SARS-CoV-2 neutralization titers (**Figure 5F-G**). This indicates that humoral immunity raised against one coronavirus is generally insufficient to generate cross-neutralizing immunity to even highly related coronavirus strains.

## DISCUSSION

Traditionally, cellular immunity is responsible for clearing an established viral infection, while humoral immune responses play a more critical role in preventing future infection. Here we found that severely ill COVID-19 patients had the highest levels of anti-RBD antibodies, which other studies have similarly described (Shrock *et al*., 2020). To further characterize this antibody response, we measured neutralization titers and developed a neutralization potency index derived from our quantitative readouts (NT50/IgG) to assess the quality of anti-RBD antibodies irrespective of the quantity produced. Remarkably, neutralization potency was significantly diminished in severely ill patients, and survival analysis demonstrated that an index of ≥100 was predictive of 100% 30-day survival, whereas <100 was associated with 87% 30-day survival in our limited cohort of 113 COVID-19 patients. Thus, this neutralization potency index may be a useful metric for physicians seeking to risk-stratify COVID-19 patients.

Despite the clear correlation between COVID-19 severity and development of humoral immunity, the cause-effect relationship between these two is unclear. One possibility is that severe disease caused by hyperinflammation and/or uncontrolled viral replication induces overproduction of antibodies that serve as a ‘biomarker’ of severity. This is supported by our finding that the most severely affected patients had the highest levels of inflammatory markers and cytokines, which can drive antibody production. In support of this possibility, a recent study suggests a pathogenic role of immune activation and exuberant antibody production from extrafollicular B cells in critically ill patients (Woodruff *et al*., 2020). Indeed, of all the COVID-19 treatment regimens being used and tested, dampening of the immune response with corticosteroids has proven to have one of the greatest benefits in improving outcomes and survival (Siemieniuk *et al*., 2020), and we find that corticosteroids decrease both anti-RBD IgG levels and neutralization titers. However, another possibility is that high levels of antibodies with low neutralization potency worsen disease severity, possibly via ADE. This is supported by our finding of decreased neutralization potency in severely ill patients, and raises concerns over the use of convalescent plasma as a treatment strategy. One exception, however, may be in immunosuppressed individuals, which generally have sub-optimal antibody levels and neutralization titers. Further studies in animal models of COVID-19 testing passive transfer of low-potency index sera may help resolve this controversy.

A multitude of vaccines are presently being evaluated for SARS-CoV-2 prevention, including inactivated virus (Gao *et al*., 2020), spike antigen (Jackson *et al*., 2020; Keech *et al*., 2020), and RBD antigen (Dai *et al*., 2020; Mulligan *et al*., 2020). Each will likely result in humoral immunity with different ratios of neutralizing and non-neutralizing antibodies. Given our results, it will be important to assess the potency index of each candidate to determine those with maximal potential. Interestingly, one study showed that vaccination of mice with RBD generated potently neutralizing antibodies without antibody-dependent enhancement. This was postulated to be due to the lack of immunodominant non-neutralizing epitopes present on the remainder of the spike protein (Quinlan *et al*., 2020).

The diverse and atypical kinetics of antibody production—in particular, early rise of IgG and in some cases IgA—suggests the possibility of a contribution from class-switched (IgG+ or IgA+) memory B cells early in the humoral immune response rather than solely from the naive (IgM+) B cell pool, as has been recently postulated (Song *et al*., 2020). Regardless, our results support a role for anti-RBD IgM and IgA in contributing to SARS-CoV-2 neutralization, despite their transient nature in serum. Anti-RBD IgG responses and neutralization, on the other hand, were sustained in the time frame analyzed (∼60 days), but several studies have emerged that question the longevity of these responses, which have yet to be determined. It is tempting to speculate that severely afflicted individuals may have more enduring immunity than mild cases. The differences in humoral response induction may stem from a combination of factors, including host permissibility to viral replication and a rapid response from innate immune effector cells and cytotoxic T cells, some of which have been postulated to arise from cross-reactive memory cells to other coronaviruses (Grifoni *et al*., 2020).

Although the mutation rate of coronaviruses is very low when compared to other viruses such as influenza or HIV, certain mutations in the spike protein of SARS-CoV-2 have emerged in the setting of the rapidly spreading pandemic. We found that one such mutation, D614G, which has now spread and become a dominant strain worldwide, does not affect the neutralizing ability of patient sera, reducing concerns for re-infection. Still, prior coronavirus pandemics (e.g. SARS-CoV, MERS-CoV, and now SARS-CoV-2) have occurred due to zoonotic coronaviruses crossing the species barrier, indicating an ongoing threat of future pandemics even in the face of effective vaccines to current viruses. One pre-emergent bat coronavirus, WIV1-CoV, is highly homologous to SARS-CoV and SARS-CoV-2 and can infect ACE2-expressing human cells (Menachery *et al*., 2016). Our data demonstrate that sera from SARS-CoV-2 infected patients exhibit very limited cross-neutralization of WIV1-CoV, except for rare individuals with relatively low-level neutralization of WIV1-CoV, suggesting that generation of broadly neutralizing antibodies is indeed possible, as has been previously described (Wec *et al*., 2020).

In summary, the development of potently neutralizing humoral immunity against SARS-CoV-2 appears to increase survival, and may protect against re-infection with other circulating strains of SARS-CoV-2. However, it is generally unlikely to provide protection against subsequent coronavirus pandemics. As such, future efforts should focus on the development of broadly active therapies and prevention modalities that generate potently neutralizing antibodies with activity across different coronavirus strains.

## Data Availability

This study assessed the diagnostic and prognostic performance of SARS-CoV-2 serological and immunological tests and was not a clinical trial. De-identified raw data of the results of serological assays can be provided upon request.

## ACKNOWLEDGEMENTS, FUNDING SUPPORT, AND DECLARATIONS OF INTEREST

We wish to thank Nir Hacohen, PhD and Jesse D. Bloom, PhD for spike-expression plasmids, and Michael Farzan, PhD for providing ACE2-expressing 293T cells. We also give thanks to Daniel Claiborne, PhD, and Vivek Naranbhai, MD, PhD, for input on experiments and statistical analyses. K.L.C. is supported by Ruth L. Kirschstein National Research Service Award (NRSA) Individual Postdoctoral Fellowship 1F32AI143480. T.M.C. and B.M.H. were supported by award Number T32GM007753 from the National Institute of General Medical Sciences. J.F. was supported by T32AI007245. A.G.S. was supported by NIH R01 AI146779 and a Massachusetts Consortium on Pathogenesis Readiness (MassCPR) grant. J.A.B. has received research support from Zeus Scientific, bioMerieux, Immunetics, Alere, DiaSorin, the Bay Area Lyme Foundation (BALF), and the National Institute of Allergy and Infectious Diseases (NIAID; Award 1R21AI119457-01) for unrelated projects. J.A.B. has served as a paid consultant to T2 Biosystems, DiaSorin and Roche Diagnostics. A.J.I. is supported by the Lambertus Family Foundation. A.B.B. is supported by the National Institutes for Drug Abuse (NIDA) Avenir New Innovator Award DP2DA040254, the MGH Transformative Scholars Program as well as funding from the Charles H. Hood Foundation. This independent research was supported by the Gilead Sciences Research Scholars Program in HIV.

## AUTHOR CONTRIBUTIONS

W.F.G.B., E.C.L., M.G.A., D.Y., T.E.M., and A.B.B. designed the experiments. W.F.G.B., E.C.L., M.G.A., D.Y., and K.L.C. carried out experiments and analyzed data. J.F., B.M.H., T.M.C., and A.G.S. provided key reagents and useful discussions and insights. D.L. offered suggestions for experiments and useful discussions. A.D.N. contributed to statistical analyses. A.J.I., J.L., J.A.B., and A.D. provided clinical samples. A.J.I., M.M., G.A. and R.C. contributed key inputs into experimental design. W.F.G.B., E.C.L., and A.B.B. wrote the paper with contributions from all authors.

## METHODS

### SARS-CoV-2 receptor binding domain IgG, IgM, and IgA ELISA

To quantitatively detect IgG, IgM, and IgA antibodies to SARS-CoV-2 receptor binding domain (RBD), we developed an indirect ELISA using an anti-SARS-CoV and -CoV-2 monoclonal antibody (CR3022) with IgG1, IgM, and IgA1 isotypes (kindly provided by Galit Alter, Stephanie Fischinger, Caroline Atyeo, and Matt Slein in collaboration with Jeffrey Bernard at MassBiologics). RBD was produced in Expi293F suspension cells with a C-terminal SBP-(His)_8_ tag followed by affinity chromatography and then size exclusion chromatography prior to removal of the His tag as described previously. 96-well Nunca MaxiSorp ELISA plates (ThermoFisher) were coated with purified RBD diluted in carbonate-bicarbonate buffer (Sigma) to a concentration of 1 μg/mL for IgG and IgA plates and 2 μg/mL for IgM plates for 1 h at room temperature. Plates were washed with a wash buffer consisting of 50 mM Tris (pH 8.0) (Sigma), 140 mM NaCl (Sigma), and 0.05% Tween-20 (Sigma). Plates were incubated with a blocking buffer consisting of 1% BSA (Seracare), 50 mM Tris (pH 8.0), and 140 mM NaCl for 30 min at room temperature, and then washed. Serum samples were diluted 1:100 with a dilution buffer consisting of 1% BSA, 50 mM Tris (pH 8.0), 140 mM NaCl, and 0.05% Tween-20. The Tween-20 detergent served as an inactivation agent to render samples non-infectious, as has been previously described for other enveloped viruses (Mayo and Beckwith, 2002). A seven-point standard curve was created using each of the standards (i.e. CR3022-IgG1, CR3022-IgM, CR3022-IgA1) starting at 2 μg/mL by performing 1:3 serial dilutions with dilution buffer. Samples and standards were added to corresponding wells and incubated for 1 h at 37°C, followed by washing. Human antibody isotypes were detected with specific antibodies (Bethyl) diluted as indicated: anti-human IgG-HRP (1:25,000), anti-human IgM-HRP (1:20,000), and anti-human IgA-HRP (1:5,000). These were added to each plate and incubated for 30 min at room temperature. After washing, TMB substrate (Inova) was added to each well and incubated for 7 min (for IgG), 13 min (for IgM), and 10 min (for IgA), before stopping with 1 M H_2_SO_4_. Buffer compositions, reagent concentrations and incubation times and temperatures were optimized in separate experiments for each analyte to maximize signal-to-noise ratio. Optical density (O.D.) was measured at 450 nm with subtraction of the O.D. at 570 nm as a reference wavelength on a SpectraMax ABS microplate reader. Anti-RBD antibody levels were calculated by interpolating onto the standard curve and correcting for sample dilution; one unit per mL (U/mL) was defined as the equivalent reactivity seen by 1 μg/mL of CR3022.

### SARS-CoV-2 pseudovirus neutralization assay

To compare the neutralizing activity of patient sera against coronaviruses, we produced lentiviral particles, pseudotyped with different spike proteins, by transient transfection of 293T cells and titered the viral supernatants by flow cytometry on 293T-ACE2 cells (Moore et al. 2004). Virus production was also quantified by p24 ELISA on viral supernatants using the HIV-1 p24^CA^ antigen capture assay (Leidos Biomedical Research, Inc). To increase throughput and consistency, assays and readouts were performed on a Fluent Automated Workstation (Tecan) using 384-well plates (Grenier). Following an initial 12-fold dilution, the liquid handler performed serial three-fold dilutions (ranging from 1:12 to 1:8,748) of each patient serum and/or purified antibody in 20 μL followed by addition of 20 μL of pseudovirus containing 125 infectious units and incubation for 1 h at room temperature. Finally, 10,000 293T-ACE2 (Moore *et al*., 2004) cells in 20 μL cell media containing 15 μg/mL polybrene were added to each well and incubated at 37°C for 60-72 h. Cells were lysed using a previously described assay buffer (Siebring-van Olst et al., 2013) and luciferase expression was quantified using a Spectramax L luminometer (Molecular Devices). Percent neutralization was determined by subtracting background luminescence measured in cell control wells (cells only) from sample wells and dividing by virus control wells (virus and cells only). Of note, repeated sera neutralization measurements in independent assays using 500, 250, 125 infectious units of pseudovirus per well generated similar results (data not shown), indicating that the NT50 is not significantly influenced by pseudovirus titers. Data was analyzed using Graphpad Prism and NT50 values were calculated by taking the inverse of the 50% inhibitory concentration value for all samples with a neutralization value of 80% or higher at the highest concentration of serum or antibody.

### Flow Cytometry

To quantify the pseudotyped lentiviral supernatants in terms of infectious units, we plated 400,000 of either 293T or 293T-ACE2 cells in 1 mL in a 12-well plate format (Corning). 24 h later, ten-fold serial dilutions of lentiviral transfection supernatant were made in 100 μL, which was then used to replace 100 μL of media on the plated cells. Cells were then incubated with lentivirus supernatant for 48 h at 37°C and then harvested with Trypsin-EDTA (Corning), resuspended in PBS supplemented with 2% FBS (PBS+), and measured on a Stratedigm S1300Exi Flow Cytometer. Samples were gated for ZsGreen expression.

To compare the relative surface expression of pseudovirus spike protein, we plated 400,000 293T cells per well in 1 mL in a 12-well plate. 24 h later, we transfected each well with a lentiviral helper vector coding for different spike proteins. The cells were incubated for 48 h at 37°C and harvested into PBS containing 1% fetal bovine serum (Sigma) (called PBS+). Cells transfected with each vector were divided into 3 aliquots, stained with either PBS+, CR3022 SARS-CoV antibody (10 μg/mL in PBS+), or B38 SARS-CoV-2 antibody (10 μg/mL in PBS+) for 30 minutes at room temperature. Cells were then washed with 1 mL PBS+, spun at 1,150 x g, and stained with anti-human IgG AF647 polyclonal antibody (Invitrogen) at 2 μg/mL in PBS+ for 30 minutes at RT. Cells were washed with 1 mL of PBS+, spun at 1,150 x g, resuspended in 150 μL of PBS+ and measured on a Stratedigm S1300Exi Flow Cytometer.

### Confocal Microscopy

60-72 hours after neutralization assay setup, each well in a serum dilution series within a 384-well plate was imaged using a FITC filter to detect cellular ZsGreen expression. Images were acquired using a 20X air objective on a Zeiss LSM510 instrument. Acquired images were analyzed using ImageJ to produce overlays.

### Patient samples

Use of patient samples for the development and validation of SARS-CoV-2 diagnostic tests was approved by Partners Institutional Review Board (protocol 2020P000895). Serum samples from 113 patients diagnosed with COVID-19 (confirmed by at least one SARS-CoV-2 PCR-positive nasopharyngeal swab at Massachusetts General Hospital) were collected over course of several weeks, resulting in partially longitudinal, cross-sectional cohort consisting of 165 serum samples, with a prospective follow-up period of at least 3 months to assess clinical course and outcomes by manual chart review by at least two physicians. For each patient, the following information was obtained: age, sex, SARS-CoV-2 PCR results, date of symptom onset, hospitalization and discharge dates, intubation and extubation dates, and deceased date. Date of symptom onset was defined as the earliest date that at least one of the following COVID-19-related symptoms was reported as developing acutely and new from baseline: fever, chills, loss of smell or taste, body aches, rhinorrhea, nasal congestion, sore throat, cough, shortness of breath. If the date of symptom onset could not be determined with confidence, this information was excluded from the analysis. Patients were assessed for the presence of absence of the following pre-existing medical conditions: lung disease (e.g. asthma, COPD), heart disease (e.g. coronary artery disease, heart failure), vascular disease (e.g. peripheral vascular disease), hypertension, diabetes, obesity (BMI >30), kidney disease, autoimmune disorder, solid organ cancer, chemotherapy for solid organ cancer, hematologic cancer, chemotherapy or immunotherapy for hematologic cancer, history of organ transplant, history of hematopoietic stem cell transplant, and pre-existing use of corticosteroids or other immunosuppressive medications. Based on these information, the cohort was divided into the following groups based on severity of disease and underlying health status: (*i*) non-hospitalized, consisting of individuals that were never admitted to the hospital and were sent home to quarantine; (*ii*) hospitalized, which included individuals that were hospitalized for at least one night but were never intubated and were eventually discharged; (*iii*) intubated, comprising hospitalized individuals that were intubated for at least one day but survived and were eventually discharged; (*iv*) deceased, for which we had obtained a specimen before they eventually passed away in the hospital; and (*v*) immunosuppressed, which consisted of people that were on immunosuppressive medication (including high-dose corticosteroid) and/or were afflicted by a clinically significant hematologic malignancy before being diagnosed with COVID-19. Laboratory data throughout admission were analyzed, and the maximum documented serum levels of ferritin, C-reactive protein, D-dimer, lactate dehydrogenase, troponin-T, and IL-6 were recorded for each patient, as well as the lowest absolute lymphocyte count documented (lymphocyte count nadir). In addition, use of the following treatments were documented: corticosteroids, hydroxychloroquine, azithromycin, atorvastatin, remdesivir, lopinavir/ritonavir, tocilizumab (part of treatment versus placebo trial, currently blinded), and anakinra. All information obtained from medical records was verified by at least two physicians.

Pre-pandemic serum samples (*n* = 1,257) were obtained from the clinicals laboratories at Massachusetts General Hospital. These samples were comprised of an unbiased cohort of individuals being tested for measles, mumps, and rubella titers (*n* = 1124), as well as a selected subset of 133 individuals with positive serology results for cytomegalovirus (*n* = 10), varicella-zoster virus (*n* = 25), hepatitis B virus (*n* = 25), hepatitis C virus (*n* = 24), HIV (*n* = 37), syphilis (*n* = 16), Toxoplasma (*n* = 1), and rheumatoid factor (*n* = 1).

### Statistical and data analyses

Statistical and data analyses were performed using GraphPad Prism 8.4.3, JMP Pro 15.0.0 (SAS Institute), and R v4.0.2. Flow cytometry data was analyzed using FlowJo 10.6.2. Non-parametric multivariate ANOVAs were performed on the indicated figures where several cohorts were present; all *p* values were adjusted for multiple comparisons except when indicated. For survival, Kaplan-Meier method was used for survival analysis, and Cox proportional hazards models performed by both JMP Pro and R confirmed these findings after accounting for additional variables. When using R, the Cox proportional hazards model was performed using the coxph function from the survival package v3.2-7 (https://CRAN.Rproject.org/package=survival) in R v4.0.2 (R Core Team 2020). In

## SUPPLEMENTARY FIGURES LEGENDS

**Supplementary Table S1: Clinical data from COVID-19 patients**. Clinical data is shown for each pre-defined severity cohort.

**Supplementary Figure S1: Clinical laboratory data from COVID-19 patients**.

(**A-D**) Violin plots of peak serum levels of (**A**) ferritin, (**B**) D-dimer, (**C**) lactate dehydrogenase, and (**D**) troponin-T documented for each COVID-19 patient in the indicated cohorts are shown. Clinical laboratory-defined cut-offs of the upper limit of normal are indicated with a dotted line. For each parameter, a non-parametric ANOVA was performed; statistical significance is indicated as follows: **** *p* < 0.0001, *** *p* < 0.001, ** *p* < 0.01, and * *p* < 0.05.

**Supplementary Figure S2: Cross-reactivity of anti-CoV antibody responses and high-throughput SARS-CoV-2 pseudovirus neutralization assay**.

**(A)** A schematic of the quantitative indirect ELISA that measures IgG, IgM, and IgA antibodies to the receptor binding domain (RBD) for SARS-CoV-2 is shown.

**(B)** Reactivity of the anti-SARS-CoV and -CoV-2-specific monoclonal antibody (CR3022 mAb) towards SARS-CoV-2, SARS-CoV, and MERS-CoV RBD was measured.

**(C)** Published crystal structures of the ACE2:prefusion-stabilized SARS-CoV-2 spike (PDB ID: 6VSB) as well as the RBD of SARS-CoV-2 (PDB ID 6VWI), SARS-CoV (PDB ID 2AJF), MERS-CoV (PDB ID: 4L72), HKU1 (PDB ID 5GNB), and NL63 (PDB ID 3KBH) are presented, with the sequence homology to SARS-CoV-2 RBD indicated.

(**D-E**) Cross-reactivity of sera from SARS-CoV-2-infected patients (*n* = 15) towards the RBD of (**D**, left) SARS-CoV and (**D**, right) MERS-CoV, as well as the reactivity of sera from healthy blood donors (*n* = 43) and COVID-19 patients towards the RBD of two common cold coronaviruses, (**E**, left) HKU1 and (**E**, right) NL63, was measured using a modified anti-RBD IgG ELISA and optical density as a read-out.

**(F)** A schematic of the full length and truncated (Δ18) construct of SARS-CoV-2 spike used to pseudotype lentivirus is shown; ERRS denotes ER retention signal.

**(G)** Expression of the indicated spike constructs was measured on the surface of 293T cells via flow cytometry.

**(H)** Pseudovirus titers of the indicated spike constructs were quantified.

**(I)** Lack of neutralizing ability of CR3022 mAb was confirmed in pseudovirus neutralization assay.

**(J)** Confocal microscopy of each well of a serum dilution series using a representative COVID-19 patient sample taken 60 - 72 h after assay setup demonstrated the correlation between luciferase activity and transduced target cells. Neutralization percentage at each dilution was calculated by measuring luciferase activity (luminescence) and normalizing to control well with no serum. Scale bar equals 200 μm.

**(K)** False positive NT50 were observed in individuals taking antiretroviral medications (*n* = 20 out of 37 individuals), while a large cohort of pre-pandemic individuals for which antiretroviral use was largely screened out showed a very low rate of infection inhibition (*n* = 12 out of 1,220).

**Supplementary Figure S3: Correlates between clinical outcomes and humoral immune responses against SARS-CoV-2**.

(**A**) Standardization of cohorts by days after symptom onset to samples collected between 14 and 42 days was done to mitigate sampling biases and balance out representation from each cohort indicated.

(**B-C**) Pearson correlations between neutralization (NT50) and (**B**) anti-RBD IgM and (**C**) anti-RBD IgA levels were performed, with *R*^2^ and *p* values indicated.

**(D)** Neutralization capacity of COVID-19 patient samples collected between 14 and 42 days after symptom onset (*n* = 85) grouped by serostatus as determined by the anti-RBD IgG, IgM, and IgA ELISA was plotted.

**(E)** Proportion of COVID-19 patients (*n* = 98) within each indicated serostatus group is presented.

**Supplementary Figure S4: Multivariate analysis of demographic data, clinical course, pre-existing medical conditions, treatments, laboratory data, and humoral immune response in COVID-19 patients**.

A multi-variate analysis of all available data including age, sex, language, hospital course and events, pre-existing medical conditions, treatments received, clinical laboratory data, and antibody and neutralization data was performed, with Pearson coefficients (*r*) ranging from −1 (red) to 0 (white) to +1 (blue). The presence of an ‘x’ indicates that there were insufficient data to correlate the variables in question. The following abbreviations were used: DASO, days after symptom onset; DAPP, days after PCR positivity; DPP, days PCR positive (total number of days between first PCR positive results and last PCR positive result that was followed by one negative result); DHos, days hospitalized; HSCT, hematopoietic stem cell transplant; CRP, C-reactive protein; LDH, lactate dehydrogenase; CK, creatine kinase; anti-RBD, anti-receptor binding domain; anti-NC Ab, anti-nucleocapsid antibody (as measured by the commercially available Roche SARS-CoV-2 total antibody chemiluminescent assay); SC2, SARS-CoV-2.

**Supplementary Figure S5. Characterization of CoV spike expression vectors**.

**(A)** Surface level expression of SARS-CoV-2 spike protein following transfection of 293T cells. Several constructs of spike were tested: codon-optimized full-length spike from SARS-CoV-2, a truncated version with 18 amino acids deleted from the cytoplasmic tail (Δ18), and a truncated version that also includes a D614G mutation. Expression was measured via flow cytometry by staining with B38 antibody at a concentration of 10 μg/mL followed by staining with an anti-human IgG antibody conjugated to AF647 at 2 μg/mL.

**(B)** Surface level expression of full-length and truncated (Δ18) WIV1-CoV spike proteins were also measured following transfection of 293T cells via flow cytometry. Expression was measured via flow cytometry by staining with CR3022 antibody at a concentration of 10 μg/mL followed by staining with an anti-human IgG antibody conjugated to AF647 at 2 μg/mL.

**(C)** Summary of expression data is shown.

(**D-E**) Titers of lentivirus pseudotyped with the (**D**) SARS-CoV-2 or (**E**) WIV1-CoV spike proteins were measured by transducing ACE2-expressing 293T cells with 100 μL of lentivirus supernatant.

(**F**) Transduction with 10-fold serial dilutions and subsequent assessment of ZsGreen expression by flow cytometry was performed to calculate pseudovirus titer (U/mL) for each construct indicated.

## REFERENCES

Arvin, A. M. et al. (2020) ‘A perspective on potential antibody-dependent enhancement of SARS-CoV-2’, Nature, 584(7821), pp. 353–363.

Beigel, J. H. et al. (2020) ‘Remdesivir for the Treatment of Covid-19 - Preliminary Report’, The New England journal of medicine. doi: 10.1056/NEJMoa2007764.

Boulware, D. R. et al. (2020) ‘A Randomized Trial of Hydroxychloroquine as Postexposure Prophylaxis for Covid-19’, The New England journal of medicine, 383(6), pp. 517–525.

Case, J. B. et al. (2020) ‘Replication-Competent Vesicular Stomatitis Virus Vaccine Vector Protects against SARS-CoV-2-Mediated Pathogenesis in Mice’, Cell host & microbe, 28(3), pp. 465–474.e4.

Chandrashekar, A. et al. (2020) ‘SARS-CoV-2 infection protects against rechallenge in rhesus macaques’, Science, 369(6505), pp. 812–817.

Chan, K. H. et al. (2005) ‘Serological responses in patients with severe acute respiratory syndrome coronavirus infection and cross-reactivity with human coronaviruses 229E, OC43, and NL63’, Clinical and diagnostic laboratory immunology, 12(11), pp. 1317–1321.

Chen, N. et al. (2020) ‘Epidemiological and clinical characteristics of 99 cases of 2019 novel coronavirus pneumonia in Wuhan, China: a descriptive study’, The Lancet, 395(10223), pp. 507–513.

Chen, X. et al. (2020) ‘Detectable serum SARS-CoV-2 viral load (RNAaemia) is closely correlated with drastically elevated interleukin 6 (IL-6) level in critically ill COVID-19 patients’, Clinical infectious diseases: an official publication of the Infectious Diseases Society of America. doi: 10.1093/cid/ciaa449.

Chen, Z. et al. (2020) ‘Efficacy of hydroxychloroquine in patients with COVID-19: results of a randomized clinical trial’, medRxiv, p. 2020.03.22.20040758.

Corver, J. et al. (2009) ‘Mutagenesis of the transmembrane domain of the SARS coronavirus spike glycoprotein: refinement of the requirements for SARS coronavirus cell entry’, Virology journal, 6, p. 230.

Crawford, K. H. D. et al. (2020) ‘Protocol and Reagents for Pseudotyping Lentiviral Particles with SARS-CoV-2 Spike Protein for Neutralization Assays’, Viruses, 12(5). doi: 10.3390/v12050513.

Dai, L. et al. (2020) ‘A Universal Design of Betacoronavirus Vaccines against COVID-19, MERS, and SARS’, Cell, 182(3), pp. 722–733.e11.

Deng, W. et al. (2020) ‘Primary exposure to SARS-CoV-2 protects against reinfection in rhesus macaques’, Science, 369(6505), pp. 818–823.

Domingo, P. et al. (2020) ‘The four horsemen of a viral Apocalypse: The pathogenesis of SARS-CoV-2 infection (COVID-19)’, EBioMedicine, 58, p. 102887.

Gao, Q. et al. (2020) ‘Development of an inactivated vaccine candidate for SARS-CoV-2’, Science, 369(6499), pp. 77–81.

Gorse, G. J. et al. (2010) ‘Prevalence of antibodies to four human coronaviruses is lower in nasal secretions than in serum’, Clinical and vaccine immunology: CVI, 17(12), pp. 1875–1880.

Grifoni, A. et al. (2020) ‘Targets of T Cell Responses to SARS-CoV-2 Coronavirus in Humans with COVID-19 Disease and Unexposed Individuals’, Cell, 181(7), pp. 1489–1501.e15.

Hannoun, C., Megas, F. and Piercy, J. (2004) ‘Immunogenicity and protective efficacy of influenza vaccination’, Virus research, 103(1-2), pp. 133–138.

Hassan, A. O. et al. (2020) ‘A SARS-CoV-2 Infection Model in Mice Demonstrates Protection by Neutralizing Antibodies’, Cell, 182(3), pp. 744–753.e4.

He, Y. et al. (2005) ‘Identification of a critical neutralization determinant of severe acute respiratory syndrome (SARS)-associated coronavirus: importance for designing SARS vaccines’, Virology, 334(1), pp. 74–82.

Horby, p. et al. (2020) ‘Dexamethasone in Hospitalized Patients with Covid-19 - Preliminary Report’, The New England journal of medicine. doi: 10.1056/NEJMoa2021436.

Iyer, A. S. et al. (2020) ‘Dynamics and significance of the antibody response to SARS-CoV-2 infection’, medRxiv : the preprint server for health sciences. doi: 10.1101/2020.07.18.20155374.

Jackson, L. A. et al. (2020) ‘An mRNA Vaccine against SARS-CoV-2 - Preliminary Report’, The New England journal of medicine. doi: 10.1056/NEJMoa2022483.

Ju, B. et al. (2020) ‘Human neutralizing antibodies elicited by SARS-CoV-2 infection’, Nature, 584(7819), pp. 115–119.

Keech, C. et al. (2020) ‘Phase 1-2 Trial of a SARS-CoV-2 Recombinant Spike Protein Nanoparticle Vaccine’, The New England journal of medicine. doi: 10.1056/NEJMoa2026920.

Kopf, M. et al. (1998) ‘Interleukin 6 influences germinal center development and antibody production via a contribution of C3 complement component’, The Journal of experimental medicine, 188(10), pp. 1895–1906.

Korber, B. et al. (2020) ‘Tracking Changes in SARS-CoV-2 Spike: Evidence that D614G Increases Infectivity of the COVID-19 Virus’, Cell, 182(4), pp. 812–827.e19.

Lemieux, J. et al. (2020) ‘Phylogenetic analysis of SARS-CoV-2 in the Boston area highlights the role of recurrent importation and superspreading events’, medRxiv : the preprint server for health sciences. doi: 10.1101/2020.08.23.20178236.

Liu, L. et al. (2019) ‘Anti-spike IgG causes severe acute lung injury by skewing macrophage responses during acute SARS-CoV infection’, JCI insight, 4(4). doi: 10.1172/jci.insight.123158.

Lontok, E., Corse, E. and Machamer, C. E. (2004) ‘Intracellular targeting signals contribute to localization of coronavirus spike proteins near the virus assembly site’, Journal of virology, 78(11), pp. 5913–5922.

Mayo, D. R. and Beckwith, W. H., 3rd (2002) ‘Inactivation of West Nile virus during serologic testing and transport’, Journal of clinical microbiology, 40(8), pp. 3044–3046.

McBride, C. E., Li, J. and Machamer, C. E. (2007) ‘The cytoplasmic tail of the severe acute respiratory syndrome coronavirus spike protein contains a novel endoplasmic reticulum retrieval signal that binds COPI and promotes interaction with membrane protein’, Journal of virology, 81(5), pp. 2418–2428.

Menachery, V. D. et al. (2016) ‘SARS-like WIV1-CoV poised for human emergence’, Proceedings of the National Academy of Sciences of the United States of America, 113(11), pp. 3048–3053.

Meng, Y. et al. (2020) ‘Sex-specific clinical characteristics and prognosis of coronavirus disease-19 infection in Wuhan, China: A retrospective study of 168 severe patients’, PLoS pathogens, 16(4), p. e1008520.

Mercado, N. B. et al. (2020) ‘Single-shot Ad26 vaccine protects against SARS-CoV-2 in rhesus macaques’, Nature. doi: 10.1038/s41586-020-2607-z.

ter Meulen, J. et al. (2006) ‘Human monoclonal antibody combination against SARS coronavirus: synergy and coverage of escape mutants’, PLoS medicine, 3(7), p. e237.

de Miranda Santos, I. K.F. and Costa, C. H. N. (2020) ‘Impact of Hydroxychloroquine on Antibody Responses to the SARS-CoV-2 Coronavirus’, Frontiers in immunology, p. 1739.

Moore, M. J. et al. (2004) ‘Retroviruses pseudotyped with the severe acute respiratory syndrome coronavirus spike protein efficiently infect cells expressing angiotensin-converting enzyme 2’, Journal of virology, 78(19), pp. 10628–10635.

Mulligan, M. J. et al. (2020) ‘Phase 1/2 study of COVID-19 RNA vaccine BNT162b1 in adults’, Nature. doi: 10.1038/s41586-020-2639-4.

Nie, J. et al. (2020) ‘Establishment and validation of a pseudovirus neutralization assay for SARS-CoV-2’, Emerging microbes & infections, 9(1), pp. 680–686.

Plotkin, S. A. (2010) ‘Correlates of protection induced by vaccination’, Clinical and vaccine immunology: CVI, 17(7), pp. 1055–1065.

Quinlan, B. D. et al. (2020) ‘The SARS-CoV-2 receptor-binding domain elicits a potent neutralizing response without antibody-dependent enhancement’. doi: 10.1101/2020.04.10.036418.

Rogers, T. F. et al. (2020) ‘Isolation of potent SARS-CoV-2 neutralizing antibodies and protection from disease in a small animal model’, Science, 369(6506), pp. 956–963.

Roy, V. et al. (2020) ‘SARS-CoV-2-specific ELISA development’, Journal of immunological methods, 484–485, p. 112832.

Shrock, E. et al. (2020) ‘Viral epitope profiling of COVID-19 patients reveals cross-reactivity and correlates of severity’, Science. doi: 10.1126/science.abd4250.

Siemieniuk, R. A. et al. (2020) ‘Drug treatments for covid-19: living systematic review and network meta-analysis’, BMJ, 370, p. m2980.

Song, G. et al. (2020) ‘Cross-reactive serum and memory B cell responses to spike protein in SARS-CoV-2 and endemic coronavirus infection’, bioRxiv : the preprint server for biology. doi: 10.1101/2020.09.22.308965.

Soresina, A. et al. (2020) ‘Two X-linked agammaglobulinemia patients develop pneumonia as COVID-19 manifestation but recover’, Pediatric allergy and immunology: official publication of the European Society of Pediatric Allergy and Immunology. doi: 10.1111/pai.13263.

Stadlbauer, D. et al. (2020) ‘SARS-CoV-2 Seroconversion in Humans: A Detailed Protocol for a Serological Assay, Antigen Production, and Test Setup’, Current protocols in microbiology, 57(1), p. e100.

Tang, W. et al. (2020) ‘Hydroxychloroquine in patients with mainly mild to moderate coronavirus disease 2019: open label, randomised controlled trial’, BMJ, 369, p. m1849.

Tersalvi, G. et al. (2020) ‘Elevated Troponin in Patients With Coronavirus Disease 2019: Possible Mechanisms’, Journal of cardiac failure, 26(6), pp. 470–475.

Tian, X. et al. (2020) ‘Potent binding of 2019 novel coronavirus spike protein by a SARS coronavirus-specific human monoclonal antibody’, Emerging microbes & infections, 9(1), pp. 382–385.

Ujike, M. et al. (2016) ‘The contribution of the cytoplasmic retrieval signal of severe acute respiratory syndrome coronavirus to intracellular accumulation of S proteins and incorporation of S protein into virus-like particles’, The Journal of general virology, 97(8), pp. 1853–1864.

Ullah, W. et al. (2020) ‘Safety and Efficacy of Hydroxychloroquine in COVID-19: A Systematic Review and Meta-Analysis’, Journal of clinical medicine research, 12(8), pp. 483–491.

Wang, L. (2020) ‘C-reactive protein levels in the early stage of COVID-19’, Medecine et maladies infectieuses, 50(4), pp. 332–334.

Wang, M. et al. (2020) ‘Remdesivir and chloroquine effectively inhibit the recently emerged novel coronavirus (2019-nCoV) in vitro’, Cell research, 30(3), pp. 269–271.

Wec, A. Z. et al. (2020) ‘Broad neutralization of SARS-related viruses by human monoclonal antibodies’, Science, 369(6504), pp. 731–736.

Woodruff, M. C. et al. (2020) ‘Extrafollicular B cell responses correlate with neutralizing antibodies and morbidity in COVID-19’, Nature immunology. doi: 10.1038/s41590-020-00814-z.

Wu, Y. et al. (2020) ‘A noncompeting pair of human neutralizing antibodies block COVID-19 virus binding to its receptor ACE2’, Science, 368(6496), pp. 1274–1278.

Wynants, L. et al. (2020) ‘Prediction models for diagnosis and prognosis of covid-19: systematic review and critical appraisal’, BMJ, 369. doi: 10.1136/bmj.m1328.

Yu, J. et al. (2020) ‘DNA vaccine protection against SARS-CoV-2 in rhesus macaques’, Science, 369(6505), pp. 806–811.

Zhou, Y. et al. (2020) ‘A New Predictor of Disease Severity in Patients with COVID-19 in Wuhan, China’, medRxiv. doi: 10.21203/rs.3.rs-29566/v1.

