## Supplementary Data for "COVID-19 neutralizing antibodies predict disease severity and survival"

### Supplementary Table 1

| Severity Cohort | non-hospitalized | hospitalized | intubated | deceased | immunosuppressed |
|---|---|---|---|---|---|
| **Total** | n = 18 | n = 45 | n = 27 | n = 10 | n = 13 |
| **Demographics** | | | | | |
| Age (median, range) | 26.5 (0 - 68) | 66 (0 - 91) | 63 (35 - 83) | 60 (39 - 77) | 60 (29 - 80) |
| Sex (% male) | 50% (9/18) | 49% (22/45) | 78% (21/27) | 70% (7/10) | 62% (8/13) |
| Language, primary (% non-English) | 39% (7/18) | 36% (16/45) | 59% (16/27) | 20% (2/10) | 8% (1/13) |
| **Clinical Course** | | | | | |
| PCR-positive days◊ (median, range) | 11 (0 - 44) | 17 (2 - 55) | 34.5 (13 - 68) | 19.5 (7 - 35) | 36 (10 - 55) |
| Hospitalized | 0% (0/18) | 100% (45/45) | 100% (27/27) | 100% (10/10) | 85% (11/13) |
| Days hospitalized (median, range) | - | 5 (1 - 41) | 43 (15 - 118) | 31.5 (1 - 66) | 10 (0 - 50) |
| Intubated | 0% (0/18) | 0% (0/45) | 100% (27/27) | 90% (9/10) | 38% (5/13) |
| Days intubated (median, range) | - | - | 26 (6 - 65) | 22.5 (0 - 57) | 0 (0 - 41) |
| ECMO | 0% (0/18) | 0% (0/45) | 7% (2/27) | 20% (2/10) | 0% (0/13) |
| Deceased | 0% (0/18) | 0% (0/45) | 0% (0/27) | 100% (10/10) | 0% (0/13) |
| **Pre-existing medical conditions** | | | | | |
| No significant conditions | 94% (17/18) | 9% (4/45) | 4% (1/27) | 20% (2/10) | 0% (0/13) |
| Lung disease | 0% (0/18) | 31% (14/45) | 11% (3/27) | 10% (1/10) | 0% (0/13) |
| Heart disease | 0% (0/18) | 36% (16/45) | 11% (3/27) | 20% (2/10) | 15% (2/13) |
| Vascular disease | 0% (0/18) | 13% (6/45) | 0% (0/27) | 10% (1/10) | 0% (0/13) |
| Hypertension | 6% (1/18) | 56% (25/45) | 70% (19/27) | 40% (4/10) | 31% (4/13) |
| Diabetes mellitus | 0% (0/18) | 40% (18/45) | 59% (16/27) | 30% (3/10) | 15% (2/13) |
| Obesity | 0% (0/18) | 22% (10/45) | 37% (10/27) | 0% (0/10) | 8% (1/13) |
| Kidney disease | 0% (0/18) | 22% (10/45) | 11% (3/27) | 10% (1/10) | 38% (5/13) |
| Autoimmune disease | 6% (1/18) | 7% (3/45) | 0% (0/27) | 10% (1/10) | 46% (6/13) |
| Hematologic malignancy | 0% (0/18) | 0% (0/45) | 0% (0/27) | 0% (0/10) | 23% (3/13) |
| Non-hematology malignancy | 0% (0/18) | 11% (5/45) | 7% (2/27) | 20% (2/10) | 0% (0/13) |
| Solid organ transplant | 0% (0/18) | 0% (0/45) | 0% (0/27) | 10% (1/10) | 46% (6/13) |
| HSC transplant | 0% (0/18) | 0% (0/45) | 0% (0/27) | 0% (0/10) | 8% (1/13) |
| Immunosuppressed | 0% (0/18) | 0% (0/45) | 0% (0/27) | 0% (0/10) | 100% (13/13) |
| **Treatments received** | | | | | |
| Hydroxychloroquine | 0% (0/18) | 9% (4/45) | 11% (3/27) | 10% (1/10) | 8% (1/13) |
| Azithromycin | 0% (0/18) | 7% (3/45) | 15% (4/27) | 10% (1/10) | 23% (3/13) |
| Remdesivir | 0% (0/18) | 7% (3/45) | 15% (4/27) | 10% (1/10) | 38% (5/13) |
| Tocilizumab† | 0% (0/18) | 22% (10/45) | 19% (5/27) | 0% (0/10) | 31% (4/13) |
| Anakinra | 0% (0/18) | 0% (0/45) | 4% (1/27) | 0% (0/10) | 0% (0/13) |
| Corticosteroids | 0% (0/18) | 7% (3/45) | 7% (2/27) | 10% (1/10) | 15% (2/13)☼ |

* PCR-positive days is defined as the numbers of days from the first documents PCR-positive result to the last document PCR-positive result that was followed by at least one PCR-negative result

† The majority of patients receiving tocilizumab were enrolled in a blinded randomized control trial with 2:1 randomization of tocilizumab to placebo, except for two patient who received it off-label

☼ In the immunosuppressed cohort, corticosteroid treatment refers to patients that newly started corticosteroids or had an increase in their baseline dose of corticosteroids (if applicable)

**Supplementary Table S1: Clinical data from COVID-19 patients.** Clinical data is shown for each pre-defined severity cohort.

#### Supplemental Figure S1

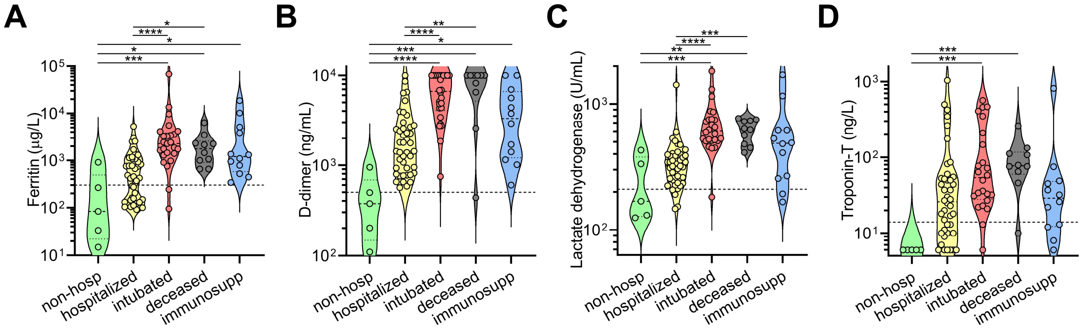


**Supplementary Figure S1: Clinical laboratory data from COVID-19 patients.**
(**A-D**) Violin plots of peak serum levels of (**A**) ferritin, (**B**) D-dimer, (**C**) lactate dehydrogenase, and (**D**) troponin-T documented for each COVID-19 patient in the indicated cohorts are shown. Clinical laboratory-defined cut-offs of the upper limit of normal are indicated with a dotted line. For each parameter, a non-parametric ANOVA was performed; statistical significance is indicated as follows: **** $p < 0.0001$, *** $p < 0.001$, ** $p < 0.01$, and * $p < 0.05$.

### Supplemental Figure S2

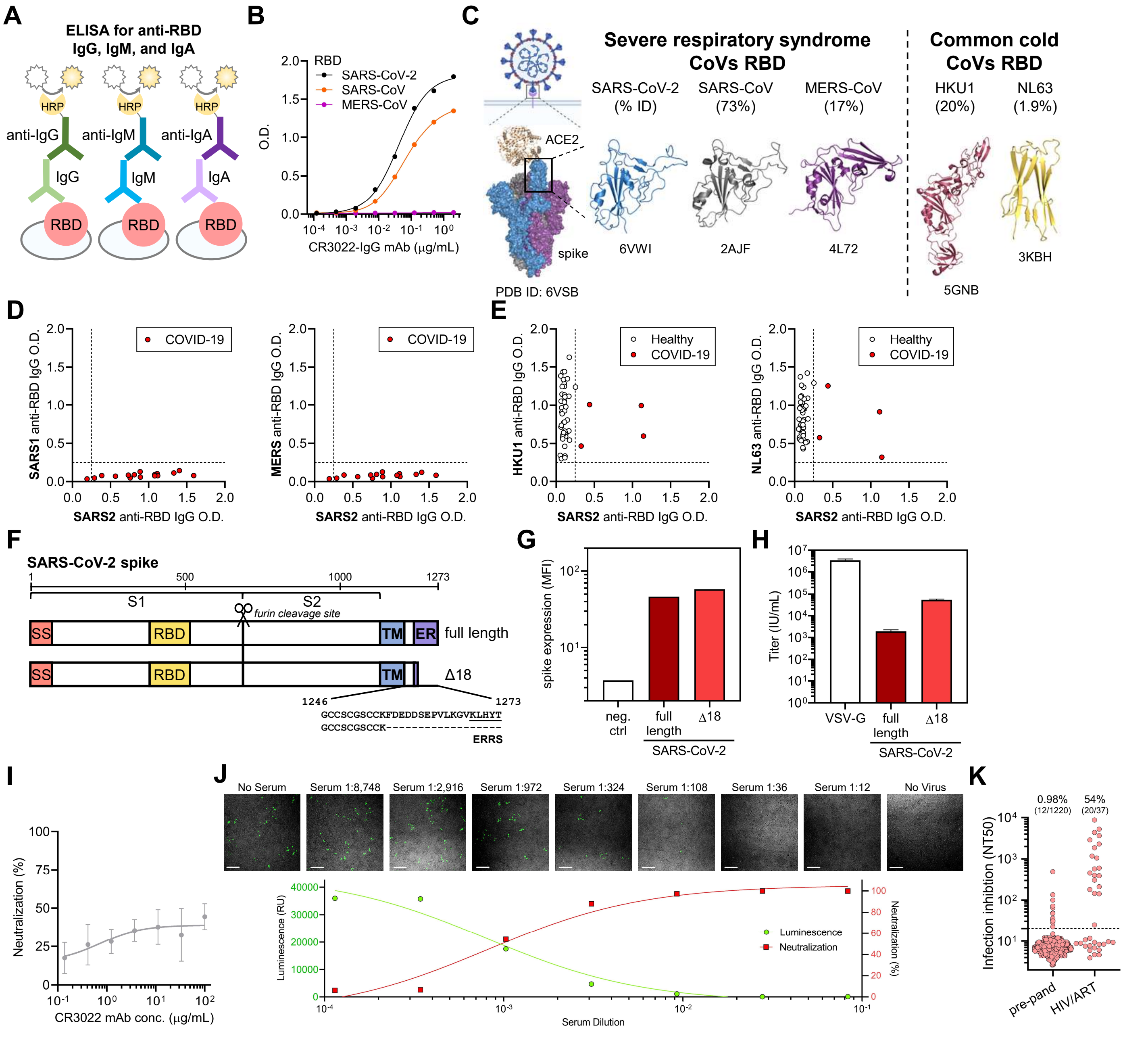


**Supplementary Figure S2: Cross-reactivity of anti-CoV antibody responses and high-throughput SARS-CoV-2 pseudovirus neutralization assay.**

(**A**) A schematic of the quantitative indirect ELISA that measures IgG, IgM, and IgA antibodies to the receptor binding domain (RBD) for SARS-CoV-2 is shown.

#### Supplemental Figure S3

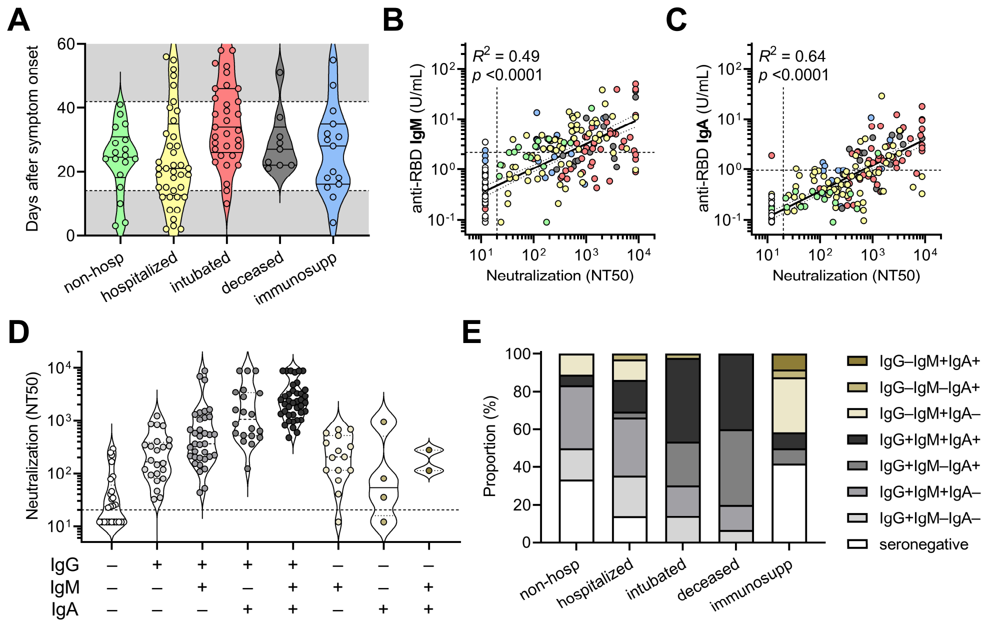


**Supplementary Figure S3: Correlates between clinical outcomes and humoral immune responses against SARS-CoV-2.**

(**A**) Standardization of cohorts by days after symptom onset to samples collected between 14 and 42 days was done to mitigate sampling biases and balance out representation from each cohort indicated.

(**B-C**) Pearson correlations between neutralization (NT50) and (**B**) anti-RBD IgM and (**C**) anti-RBD IgA levels were performed, with $R^2$ and $p$ values indicated.

(**D**) Neutralization capacity of COVID-19 patient samples collected between 14 and 42 days after symptom onset ($n$ = 85) grouped by serostatus as determined by the anti-RBD IgG, IgM, and IgA ELISA was plotted.

(**E**) Proportion of COVID-19 patients ($n$ = 98) within each indicated serostatus group is presented.

#### Supplemental Figure S4

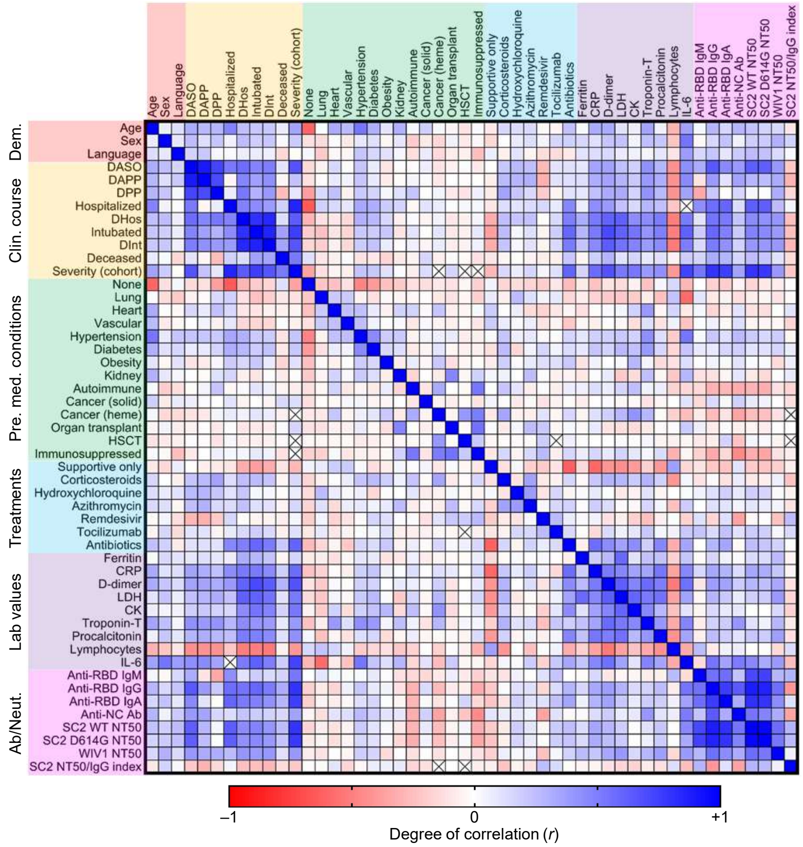


**Supplementary Figure S4: Multivariate analysis of demographic data, clinical course, pre-existing medical conditions, treatments, laboratory data, and humoral immune response in COVID-19 patients.**

A multi-variate analysis of all available data including age, sex, language, hospital course and events, pre-existing medical conditions, treatments received, clinical laboratory data, and antibody and neutralization data was performed, with Pearson coefficients (*r*) ranging from -1 (red) to 0 (white) to +1 (blue). The presence of an 'x' indicates that there were insufficient data to correlate the variables in question. The following abbreviations were used: DASO, days after symptom onset; DAPP, days after PCR positivity; DPP, days PCR positive (total number of days between first PCR positive results and last PCR positive result that was followed by one negative result); DHos, days hospitalized; HSCT, hematopoietic stem cell transplant; CRP, C-reactive protein; LDH, lactate dehydrogenase; CK, creatine kinase; anti-RBD, anti-receptor binding domain; anti-NC Ab, anti-nucleocapsid antibody (as measured by the commercially available Roche SARS-CoV-2 total antibody chemiluminescent assay); SC2, SARS-CoV-2.

#### Supplemental Figure S5

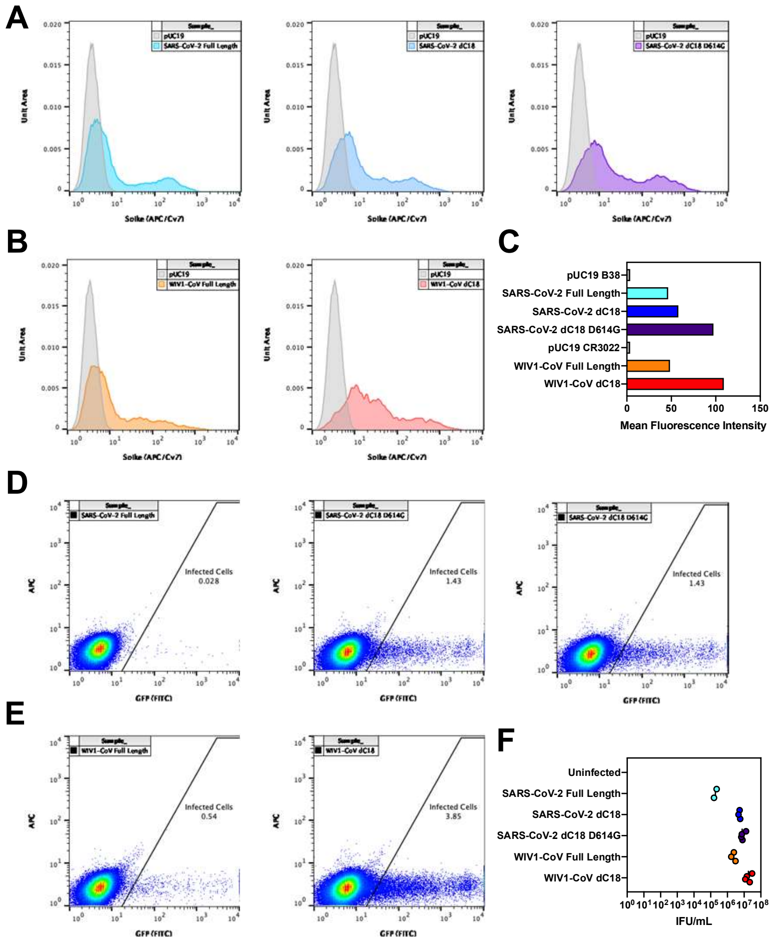


**Supplementary Figure S5. Characterization of CoV spike expression vectors.**

(**A**) Surface level expression of SARS-CoV-2 spike protein following transfection of 293T cells. Several constructs of spike were tested: codon-optimized full-length spike from SARS-CoV-2, a truncated version with 18 amino acids deleted from the cytoplasmic tail (Δ18), and a truncated version that also includes a D614G mutation. Expression was measured via flow cytometry by staining with B38 antibody at a concentration of 10 µg/mL followed by staining with an anti-human IgG antibody conjugated to AF647 at 2 µg/mL.

(**B**) Surface level expression of full-length and truncated (Δ18) WIV1-CoV spike proteins were also measured following transfection of 293T cells via flow cytometry. Expression was measured via flow cytometry by staining with CR3022 antibody at a concentration of 10 µg/mL followed by staining with an anti-human IgG antibody conjugated to AF647 at 2 µg/mL.

(**C**) Summary of expression data is shown.

(**D-E**) Titers of lentivirus pseudotyped with the (**D**) SARS-CoV-2 or (**E**) WIV1-CoV spike proteins were measured by transducing ACE2-expressing 293T cells with 100 µL of lentivirus supernatant.

(**F**) Transduction with 10-fold serial dilutions and subsequent assessment of ZsGreen expression by flow cytometry was performed to calculate pseudovirus titer (U/mL) for each construct indicated.
